# National-scale surveillance of emerging SARS-CoV-2 variants in wastewater

**DOI:** 10.1101/2022.01.14.21267633

**Authors:** Fabian Amman, Rudolf Markt, Lukas Endler, Sebastian Hupfauf, Benedikt Agerer, Anna Schedl, Lukas Richter, Melanie Zechmeister, Martin Bicher, Georg Heiler, Petr Triska, Matthew Thornton, Thomas Penz, Martin Senekowitsch, Jan Laine, Zsofia Keszei, Beatrice Daleiden, Martin Steinlechner, Harald Niederstätter, Christoph Scheffknecht, Gunther Vogl, Günther Weichlinger, Andreas Wagner, Katarzyna Slipko, Amandine Masseron, Elena Radu, Franz Allerberger, Niki Popper, Christoph Bock, Daniela Schmid, Herbert Oberacher, Norbert Kreuzinger, Heribert Insam, Andreas Bergthaler

## Abstract

SARS-CoV-2 surveillance is crucial to identify variants with altered epidemiological properties. Wastewater-based epidemiology (WBE) provides an unbiased and complementary approach to sequencing individual cases. Yet, national WBE surveillance programs have not been widely implemented and data analyses remain challenging.

We deep-sequenced 2,093 wastewater samples representing 95 municipal catchments, covering >57% of Austria’s population, from December 2020 to September 2021. Our <u>Va</u>riant <u>Qu</u>antification in S<u>e</u>wage pipeline designed for <u>Ro</u>bustness (*VaQuERo*) enabled us to deduce variant abundance from complex wastewater samples and delineate the spatiotemporal dynamics of the dominant Alpha and Delta variants as well as regional clusters of other variants of concern. These results were cross validated by epidemiological records of >130,000 individual cases. Finally, we provide a framework to predict emerging variants *de novo* and infer variant-specific reproduction numbers from wastewater.

This study demonstrates the power of national-scale WBE to support public health and promises particular value for countries without dense individual monitoring.

**Graphical Abstract:** 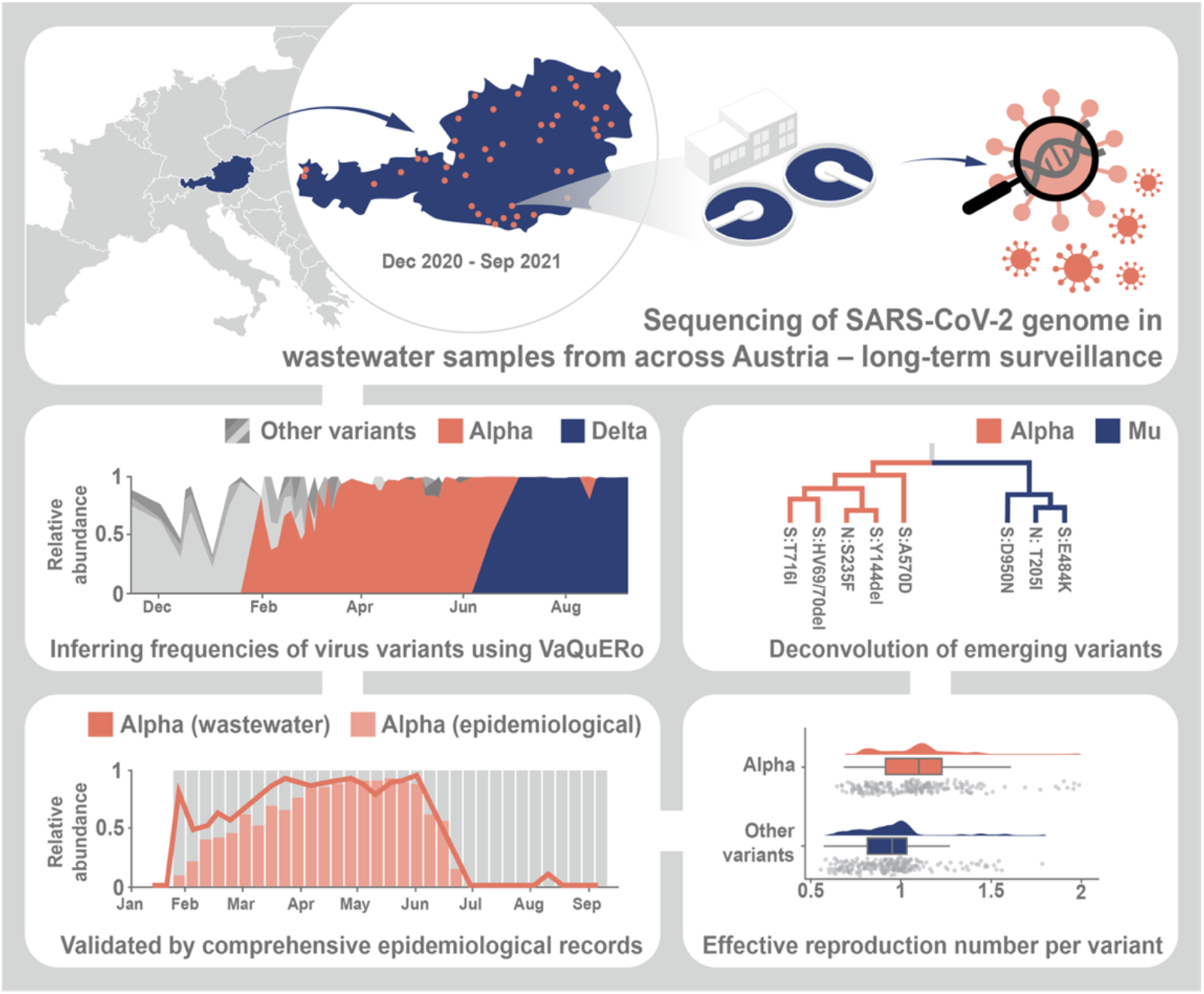

## Introduction

The recent introduction of SARS-CoV-2 into the human population led to a pandemic with immense health and socioeconomic impact worldwide ^1,2^. The pace of the successful development and use of protective vaccines has been unprecedented and is widely considered to be instrumental to end the pandemic ^3,4^. Yet, the emergence of new variants with higher transmissibility and/or immune escape properties may pose future challenges for containing the circulation of the virus ^5,6^. The evolution of SARS-CoV-2 is widely studied but remains incompletely understood ^7,8^. Importantly, potential contributing factors like antiviral immunity by natural infections or vaccines as well as social behaviour, reservoirs such as long-term infected immunosuppressed patients and animals, testing strategies and global transmission make it difficult to predict the dynamic nature of the future evolution of SARS-CoV-2 ^9–12^.

More than twenty months into the pandemic, the importance of virus surveillance for public health continues to increase worldwide. Typically, this requires the identification of SARS-CoV-2 positive cases and the underlying viral genome signature by either mutation-specific polymerase chain reaction (PCR) or sequencing-based methods. Comprehensive SARS-CoV-2 surveillance programs allow to rapidly monitor the epidemiological situation and link it to viral variants and clinical outcomes ^13,14^. Results thereof are of pivotal importance for decision makers to assess the current situations and prepare for imminent developments within but also beyond the respective countries ^13^. Nonetheless, such programs do come with limitations in their applicability on a global scale. Certain population groups including asymptomatic and underprivileged individuals may be inherently underrepresented depending on the given national testing and surveillance strategy in place ^15^. Furthermore, few countries can muster enough resources for such comprehensive monitoring, which includes adequately funded organizational structures, cross-disciplinary scientific expertise, and the readiness to integrate these resources into public health operations ^16–18^.

Wastewater-based epidemiology (WBE) promises to overcome sampling bias and economic constraints of epidemiological case-based monitoring programs ^19^. SARS-CoV-2 is excreted by more than half of infected patients via faeces ^20–22^. Additionally, sputum, saliva and urine are also discharged into the sewage system and contribute to the virion load in the wastewater (WW). Hence, municipal WW drainage systems can serve as point of use for representative sampling of circulating SARS-CoV-2 variants in a qualitative and quantitative manner ^23,24^. Initially, refined PCR-based approaches were applied to detect, at times even retrospectively, viral RNA in the sewage ^25–30^. Deduced prevalence was shown to empower forecasts of infection dynamics for the near future ^31^. The successful sequencing of SARS-CoV-2 genomes from WW was reported ^32,33^ and used to detect regional occurrence of selected virus variants based on the presence of selected characteristic mutations. ^26,34–48^. Later, WW was used to construct and quantify haplotype signatures of variants of concern and to deduce the reproductive number ^49,50^. Yet, the suitability of WBE to survey the spatiotemporal viral dynamics and integrate variant-specific epidemiological parameters on a national-scale was not assessed so far.

In Austria, a Central European country of around 9 million inhabitants surrounded by eight countries and with a high degree of international mobility due to tourism and external economic ties, the first pandemic wave from February to April 2020 was curbed by strict lockdown measures (**Extended Data Fig. 1**) ^51–53^. This coincided with early efforts to establish a viral surveillance system in WW in Austria. In March 2020, a broader surveillance approach was successively implemented. Since December 2020, close to the peak of the second wave, up to 95 wastewater treatment plants (WWTP) were sampled on a weekly basis, covering >57% of the Austrian population (**Extended Data Fig. 2**). Like in other countries in Europe and beyond, the Alpha variant (pangolin nomenclature ^54^: B.1.1.7) was introduced in December 2020, which was followed by a near complete take-over by the Delta variant (pangolin nomenclature: B.1.617.2 and AY.*) by August 2021 ^55^. Notably, the Austrian province of Tyrol harboured one of the largest clusters of the Beta (B.1.351) variant outside of South Africa from January to March 2021 ^56^. Such spatiotemporal dynamics of emerging viral variants coupled with a national epidemiological surveillance system ^57^, including comprehensive and routine genotyping of positive cases, provided the stage to develop and validate the necessary methodology for WW-based variant monitoring by sequencing.

In this study, we developed and validated a robust framework to deduce public health-relevant epidemiological indicators such as relative variant abundance and variant specific reproduction numbers from WW-derived deep sequenced SARS-CoV-2 genomes within the context of a national-scale WBE program.

## Results

### Data collection

For the presented study, samples from selected WWTPs across Austria, collected between December 1, 2020, and September 15, 2021, were considered. All samples were subjected to RT-qPCR based measurements of SARS-CoV-2 genome quantity, using internal spike-in controls to deduce absolute copy number per volume. Wastewater collection, pre-processing and quantitative screening was performed at three different laboratories. Thereof, a total of 2,093 representative samples from 95 different WWTP across Austria were further analysed by amplicon-based whole genome deep sequencing (**Fig. 1a**). The selected WWTP varied in size between 1,490 to 1,900,000 connected persons (median size: 22,370). Associated catchment areas included 57.3% of the total Austrian population. Obtained sequencing data were analysed with a PCR amplicon-based deep sequencing pipeline ^52^. Of all samples 14.7% failed during sequencing, most of the time due to low viral load in the WW as indicated by C_T_ values >35. The remaining 85.3% samples exhibited a convincing detection of SARS-CoV-2, but only 70.7% passed our quality criteria of at least 40% genome coverage, to be considered for follow up analysis (**Extended Data Fig. 2**).

**Fig. 1.**
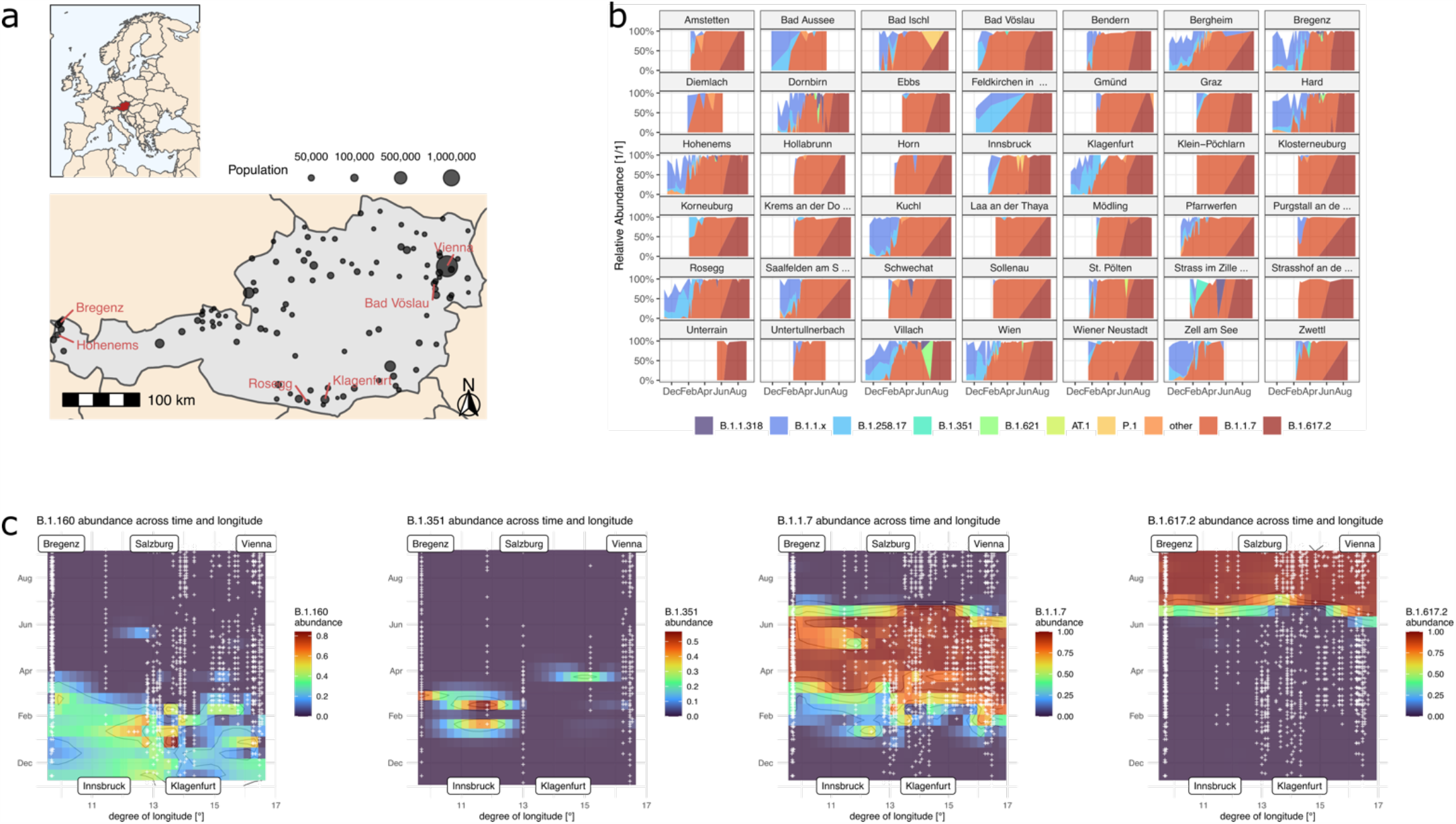
Variant Quantification. **a**, Location of Austrian WWTP considered. WWTP labelled with names are used to showcase analysis details throughout the manuscript. **b**, Results of the *VaQuERo* variant quantification analysis for the ten most abundant variants for all WWTP with more than 10 timepoints. **c**, Spatiotemporal spread of selected variants. Time is projected on the vertical axis, longitude is projected on the horizontal axis, colour indicates relative variant abundance, B.1.160, Beta (B.1.351), Alpha (B.1.1.7), Delta (B.1.617.2), from left to right, as deduced from the measured points and interpolated with B-splines. White crosses indicate measured data points.

The reproducibility of our sequencing pipeline to call low-frequency mutations down to 1% in clinical samples was shown previously ^52^. To validate mutation calling in wastewater samples, we sequenced triplicate samples from two representative locations, WWTP Hofsteig with a high industrial share and more rural WWTP Kuchl. Comparing the observed mutations qualitatively between the triplicates revealed that only 29% and 21% of the mutations in the WWTPs Hofsteig and Kuchl, respectively, were observed in all three replicates (**Extended Data Fig. 3a**). For mutations called by at least two of the three replicates this number rose to 47% and 40%, respectively. Despite the high dropout rate, the overall correlation of observed mutation frequencies was encouraging with correlation factor in pairwise comparisons between 0.69 and 0.87 (**Extended Data Fig. 3b**). Notably, the read coverage along the genome deviated from a uniform distribution, leaving substantial regions with only a few informative reads. The positioning of lowly covered regions did not show a strong reproducibility between the replicas, indicating that variable sample quality and stochastic reason have a more pronounced effect, than systematic technical causes, such as amplicon primer affinities (**Extended Data Fig. 3c**). Consequently, mutations which were expected to be present, e.g. defining mutations for a viral strain dominating at a certain time, often failed to manifest in the data (**Extended Data Fig. 3d**). Comparing the deduced mutation frequencies from the single samples to the median value across the respective triplicate also revealed substantial deviation from the expected mutation frequency, especially in the range up to 50% relative abundance (**Extended Data Fig. 3e**). Most missed mutations were in regions of low read coverage which explains their dropout. However, some of the missed mutations would have been covered by a substantial number of reads. This lack of detection may be owed to sampling errors during the amplification step due to the overall low amounts of viral RNA in the complex WW matrix (**Extended Data Fig. 3d**,**e**).

### Variant characterization and quantification

Our technical assessments of WW sequencing determined the overall reliability and reproducibility of the approach, but also showed inherent shortcomings related to dropouts at individual sites. Consequently, we aimed to develop a robust and error-tolerant method for the detection and quantification of different viral variants, which we named *VaQuERo* (<u>Va</u>riant <u>Qu</u>antification in S<u>e</u>wage designed for <u>Ro</u>bustness). To increase the positive predictive value, a reduced set of relevant variants for targeted quantification is defined first. Thereby, detectable variants are restricted to all variants defined as variants of concern, of interest or under monitoring, as defined by the European Centre for Disease Prevention and Control (ECDC), and variants circulating in Austria according to GISAID database records ^58,59^. In total 36 variants of relevance were included (**Extended Data Fig. 4**). Each mutation which occurred in more than 85% of all genomes of a particular variant deposited in the GISAID database, was considered a marker mutation for the respective variant. Mutations found in only one of the variants of relevance were denominated as unique markers (**Extended Data Fig. 4**). Based on this comprehensive set of marker mutations, we developed a method to detect and quantify variants. To gain robustness against the anticipated dropouts and hence inflated number of mutations with an observed frequency of zero, we implemented a hurdle model inspired scheme ^60^. In a first step, we removed all mutations not found in the set of the marker mutations or that were observed at a mutation frequency below 2% ^61^. Variants of relevance, with at least three unique mutations remaining, were designated as detected and subjected to the subsequent quantification step. For quantification both unique and non-unique marker mutations were used in a SIMPLEX regression, to deduce the overall variant frequencies of the detected variants of relevance. Applying *VaQuERo* to our national-scale WW data enabled us to quantify the temporal development of the variant composition for each WWTP, and hence for its contributing population in the sewer-catchment area (**Fig. 1b**). Thereby, the *VaQuERo* approach converted the detailed but complex mutation patterns as observed in the WW into a format that was reminiscent of infection case-based epidemiological reports and made it more accessible for public health stakeholders in Austria. Moreover, as these data provides information on variant-specific relative abundances over time, it can also serve as the basis for further in-depth analysis, e.g. analysis of differential fitness of distinct variants (**Extended Data Fig. 5**).

To visualize the regional expansion and decline of single variants, we reduced the Austrian geographical drawn-out shape (West-East: 575 km [7.6°], North-South: 294 km [2.6°]) to its longitudinal dimension and supplemented it by the factor time as an additional dimension (**Fig. 1c**). Interpolation in this reduced space-time coordinate system allowed to illustrate the spatiotemporal development of variant incidence. This was demonstrated by the steady retreat of B.1.160 and its replacement by Alpha, ushering in the eastern provinces of Austria during January and being completed in the western provinces only around three months later. In contrast, the exchange of Alpha by the variant B1.617.2 started almost concomitantly throughout Austria and was already completed (>98%) a month later during July 2021 (**Fig. 1c**).

### Validation of the results

In a next step, we sought to validate our WBE analysis results through a comprehensive integration with available epidemiological data from individual cases in the catchment areas (> 130,000) collected by the Austrian Agency of Health and Food Safety (AGES). Noteworthy, during the period from Jan 1^st^ and Sept 15^th^, 2021, 70.8% of all positive cases in Austria were examined if they constitute a variant of concern. Together with the overall high testing rate and the low positive rate ^57^, the number of unreported cases is assumed to be comparatively low. The absolute case counts per detected variants from communities within the catchment areas, were aggregated to proportions per total genotyped cases per week per catchment area. Contrasting the two independent data sources demonstrated a high concordance of the WW data analysis with the epidemiological trends and the time of introduction of to-be dominant variants (**Fig. 2a**). WBE also reliably captured the detection of smaller epidemiologically confirmed outbreaks as shown, for example, for Beta (pangolin nomenclatur: B.1.351) and Mu (pangolin nomenclature: B.1.621) in the municipal area discharging to the WWTP Bregenz (**Fig. 2a**, top left). Occasional flare-ups were detected in the WW even with only a marginal number of positively tested individuals residing in the respective catchment area. In some instances, such as the Mu variant in WWTP Hohenems (population: 43,722) or B.1.318 in WWTP Rosegg (population: 23,600), even timepoints with a single confirmed infection case in the catchment area were reflected by a proportionate signal in the WW analysis (**Fig. 2a**). Collectively, the signal from the WW variant surveillance and the epidemiological case monitoring correlated with an R^2^ of 0.73 (**Fig. 2b**). To estimate the sensitivity of the WW based approach, the signal from the epidemiological case monitoring program was examined for regions and timepoints where no corresponding variant-specific signal in the WW surveillance was detected. These missed variant occurrences constituted between 1 and 46 epidemiologically assigned cases, with a median of just 2 cases. In terms of relative frequencies, missed variant signals range from 0.02% to 100% with a median value of 2.38%, which is close to the expected detection limit of our method, given that mutations with frequencies below 2% in wastewater samples are dismissed as not reliable in the first place (**Fig. 2c**). Considering the median missed case count and the median missed variant frequency altogether, 81% of all missed datapoints were below either of them, indicating an estimated effective detection limit of the applied method. Conversely, a variant linked to more than 2 cases and more than 2.38% of all cases in the catchment area can be expected to be reliably detected in the WW. Overall, the comparative analyses revealed, that WBE provides a detailed reflection of the dynamic epidemiological situation down to the regional level.

**Fig. 2.**
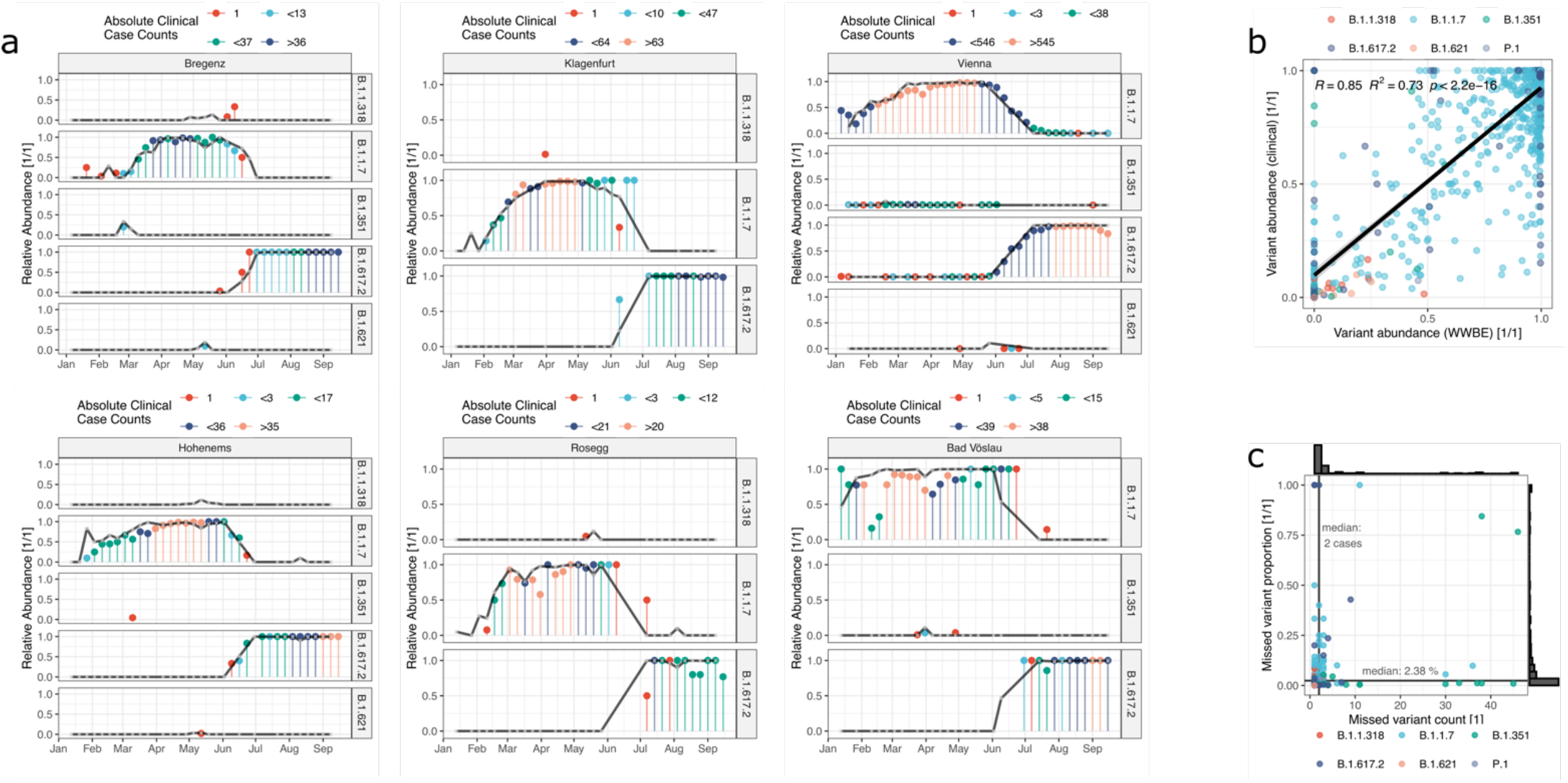
Validation. Comparison between relative variant abundance as deduced from WW variant surveillance and from the aggregated statistics of the epidemiological case-based variant monitoring program by the Austrian Agency for Health and Food Safety. Data are aggregated over a week and catchment areas. **a**, Comparison in detail for a selection of six WWTP from west, central and eastern Austria, for each a small and large population size are selected. Black line represents the signal from the WW. The capped bars (lollipop plot) represent the results of the epidemiological monitoring program, whereas the respective colour indicates the range of absolute case numbers for the respective catchment area and week. The chosen breaks correspond to the 0, 0.2, 0.5, 0.75 quantile of the observed zero-truncated absolute case count distribution. **b**, depicting the overall correlation across all detected variants and all surveyed WWTP. **c**, depicting the absolute case counts and the relative case count for all data points for which no corresponding variant specific signal was detected in the WW. Black bars and depict distribution for both variables, black line the corresponding median value.

### De-novo recognition of undescribed mutation constellation

Beside the reliable detection and relative quantification of known variants, detecting novel emerging variants is the second prospect a variant surveillance program should serve. Effectively, this is equivalent to a deconvolution of single haplotypes in the amalgamation of genotypes found in the sewage. Our amplicon-based sequencing approach relies on a modified version of the widely used ARTIC primer set and amplicons of around 400 bases ^62^, which renders direct linkages of mutations across amplicons largely unfeasible. To overcome this obstacle, we conceived to conflate associated mutations by their corresponding frequency pattern. Thereby, it was possible to designate different mutation constellations based on their observed frequencies across different samples in time and space by a hierarchical, unsupervised two-step clustering approach (see Methods). The produced constellations of correlated mutations were characterized by incorporating them into the SARS-CoV-2 reference genome with subsequent variant typing with the software tool pangolin ^54^. Evaluation of this approach showed that single samples typically feature not enough supportive information to deduce telling mutation constellations. Apparently, a sensible composition of the sample set is pivotal to produce reliable results. In this line, our de novo mutation constellation detection approach was applied to all samples from the same calendar week and the same federal state. To demonstrate at what point in time the emergence of an Alpha typic mutation constellation could have been identified, we exemplarily investigated WWTP from the province of Carinthia, for which dense sequencing data well before the arrival of Alpha is available. With our approach, Alpha-related mutations were observed in a dedicated constellation deduced from six samples from February 1^st^ to February 6^th^, 2021 (calendar week 5). The respective constellation contained a total of 30 mutations, 19 of which are defining mutations for Alpha (**Fig. 3a**). To evaluate the relevance of the obtained mutation constellations, this way determined mutation can again be used as input for *VaQuERo* to examine the timely trends and geographic spread of the de novo identified mutation constellations. Such an analysis immediately displays that the observed constellation is indeed novel and sharply rising in three out of four WWTP (**Extended Data Fig. 6**).

**Fig. 3.**
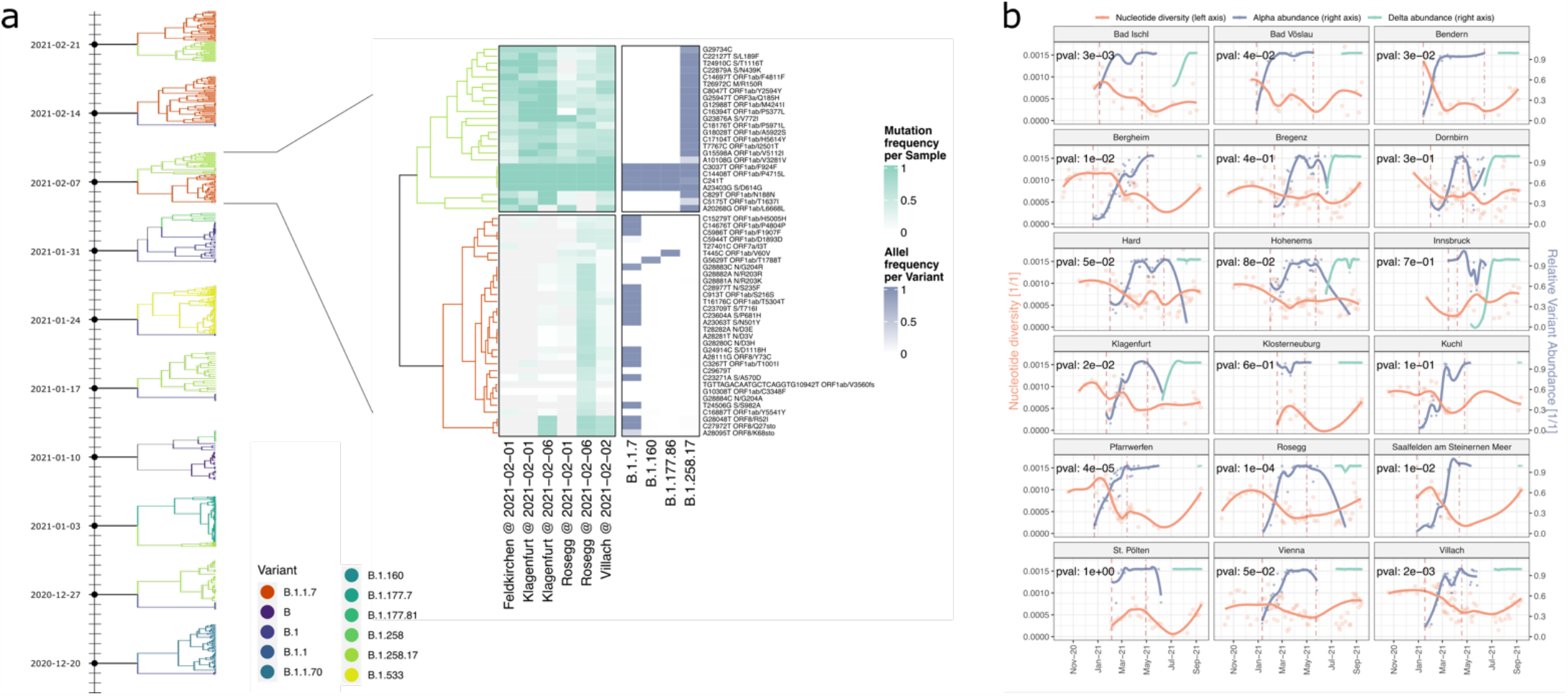
Analysis of mutation patterns. **a**, Mutation constellations in tree representation (left side) as agglomerate according to their mutation frequencies. Samples from Carinthia were jointly analysed per sampling week in the time period from December 2020 to February 2021. The first constellation enriched with Alpha (B.1.1.7) mutations is showcased with the frequencies of the mutations as observed in the samples used for deconvolution (heatmap, left side) and the frequency of the respective mutation in all samples of a specific variant as deposited in GISAID (heatmap, right side). Mutations of both constellations were traced over time using *VaQuERo* (**Extended Data Fig. 6**). **b**, Comparison of the relative abundance of Alpha (B.1.1.7) and Delta (B.1.617.2) and the observed nucleotide diversity π in the WW samples. Time course data smoothed by locally weighted regression (loess). Upper left corner indicates p-value that nucleotide diversity is higher around the first observation of Alpha (B.1.1.7) (dashed vertical line) compared to timepoint of maximal Alpha (B.1.1.7) abundance (dot-dashed vertical line) based on a Mann-Whitney U test.

### Nucleotide Diversity

One important concept in molecular population biology is the nucleotide diversity π, expressing the mean number of nucleotide differences across all loci between two genome sequences from all possible pairs ^63^. A small π indicates a very homogeneous population. Emerging variants with a higher reproductive fitness, gradually outdistancing prevailing variants, are expected to concomitantly reduce observed π-values. In contrast, repeated introduction of virus through infected individuals should manifest in an elevated π-value. To test these hypotheses, we first calculated the nucleotide diversity π for each sequenced sample and compared its progression with the relative amount of the Alpha and Delta variants, respectively (**Fig. 3b**). The recurring pattern seen, is an increasing or stable nucleotide diversity π prior to the introduction of Alpha. After Alpha had arrived and became the dominant variant, the level of π was significantly reduced in the majority of WWTPs (**Fig. 3b**). This reduction, which even coincides with an increase of prevalence, demonstrates the ongoing selective sweep in the genomic diversity due to Alpha replacing previously prevalent variants (**Fig. 3b**). In contrast, upon the abrupt takeover of the Delta variant no further reduction of π can be observed (**Fig. 3b**). Since the effect of the selective sweep during the Alpha transition dominates the nucleotide diversity in the measured time period, further hypotheses were subsequently evaluated only on samples with at least 95% relative frequency of the Alpha variant. Population size and number of active infections in the catchment area positively correlated with observed nucleotide diversity (**Extended Data Fig. 7a**,**b**). Yet, this explained only a small fraction of the observed variability (R^2^ of 0.03). To test the hypothesis that regional migration of persons contributed to elevated π-values, we resorted to time resolved mobile communication network records and official numbers of Austria’s Federal Statistical Office on registered commuters and registered touristic overnight stays. None of these mobility-based analyses could confirm a correlation between people’s regional movements and diversity of the virus population measured in the sewage (**Extended Data Fig. 7c-f**).

### Deduction of a variant specific reproduction number from wastewater

Sequencing-based quantification typically produces only relative, and not absolute quantification of single variants. Yet, it can be combined with ex-ante measured RT-qPCR based quantification of the total viral load of the same sample ^50^. Thereby, it can be assumed that the SARS-CoV-2 load normalized per population equivalents λ reflects a proxy of the prevalence of COVID-19 within the population connected to a sewage-system ^64^. Accordingly, its first derivative with respect to time dλ_*v*_/dt is a surrogate for the effective reproduction number R_eff_ ^65^, which we denoted as reproduction number from wastewater R_ww_. Hence, our data allowed a time-, region-, and variant-resolved tracing of the effective reproduction number (R_ww_), an important epidemiological parameter (**Extended Data Fig. 8**). Variant specific R_ww_ values were calculated considering the total viral load according to RT-qPCR normalized to the ammonium-nitrogen load as population size marker and the variant specific relative abundances derived by *VaQuERo* from the same WW sample. R_ww_ of Delta could not be deduced this way since the rapid spread of Delta during a low incidence phase did not produce enough timepoints with a signal of Delta and a concomitant signal of a competing variant. For the variant of concern Alpha, the variant specific R_ww_ was increased by a factor 1.34 compared to concomitantly circulating variants. This result was robustly confirmed in all examined WWTP (**Fig. 4**) and is supported by epidemiological studies ^66–68^.

**Fig. 4.**
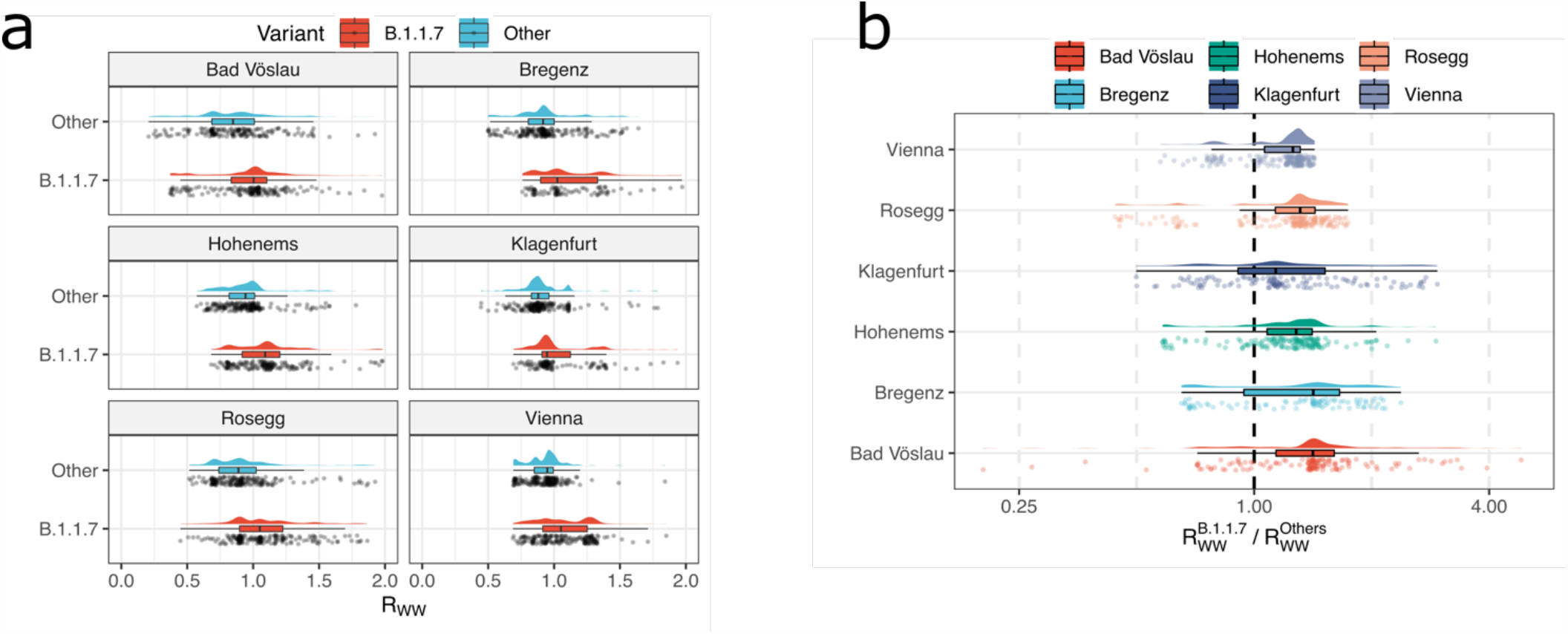
Quantitative trend analysis of virus load interlaced with variant quantification. **a**, Distribution of R_ww_ for the variant Alpha (B.1.1.7) and all other variants as calculated by an integration of absolute case estimate by RT-qPCR and variant estimate by sequencing, showcased for six WWTP across the time period from January to June, 2021. **b**, The ratios of the variant specific R_ww_ from the same timepoint and the same WWTP exhibit a systematic shift to the values >1, with a median value of 1.33, indicating an 33% increased transmissibility of Alpha (B.1.1.7) over other competing variants subjected to equal environment.

## Discussion

Foremost, we aimed with this study to convert insights deduced from WBE into classical, established concepts of case-based SARS-CoV-2 epidemiology. This includes the detection and quantification of a broad range of variants as defined by pangolin and the calculation of WW derived reproduction number (R_ww_). Thereby, we facilitate its direct utilization in decision making processes of public health authorities. We demonstrated that our newly developed *VaQuERo* approach allows to robustly deduce relative virus variant frequencies from WW. For the first time it could be shown in large-scale that variant surveillance from WBE and case-based epidemiology agree qualitatively and quantitatively. Onset, duration and scale of variant prevalence are in good agreement. The robustness of this finding is warranted by the comprehensive testing strategy implemented in Austria, and the longitudinal and transversal breadth of the here presented and publicly shared WW sequencing data. With respect to detection sensitivity of single cases, we note that the accuracy of sequencing based WBE depends on prevalence and population number in the catchment area. In a catchment area with too many positive cases, the signal of a particular case becomes undetectable in the overall signal. Too low of a prevalence rate leads to a high dilution of the virions in the WW, thus impedes the detection of the overall signal. On average, we observed with the given relation of (time variable) prevalence and catchment size, that an absolute signal >2 cases or a relative signal >2.38% could be reliably identified. Thus, the presented method may prove helpful to survey and identify new viral variants in an early stage after regional introduction.

Methods for the detection of de novo variants from the complex genotype mixtures in the WW are still in their infancy. Our approach to assort mutations based on their individual mutation frequencies across several samples, serves as a first proof of principle and can be integrated directly with the variant definition guided *VaQuERo* approach to identify emerging mutation constellations, and investigate their temporal and spatial development patterns. A robust confirmation that the deduced constellations indeed are novel haplotypes remains to be shown based on individual patient samples.

In contrast to clinical samples, virus sequencing from WW remains a technically challenging undertaking due to the overall low concentration of target molecules as well as numerous interfering factors in the complex WW matrix. The former can be dealt with using appropriate enrichment strategies while the latter may be more difficult to control for. The manifold possible combinations between the WW matrix influencing factors (e.g. seasonal changes, municipal versus industrial discharger), sewage system architecture (e.g. stormwater implementation), sample pre-processing and sequencing techniques, still await a thorough investigation for a detailed understanding of interferences and protocol adaptations. It is important to highlight that the here presented results were accomplish despite considerable deficiencies in the raw sequencing data quality caused by the complex wastewater matrix, making it applicable to versatile operating conditions. However, fine-grained sample designs ^69^ and/or more customized extraction methods ^70^ may promise an even higher resolution of variant designation, detection and quantification for appropriate settings.

Our results indicate that WBE recapitulates epidemiological screening programs at a high spatiotemporal resolution, at reduced sample number and logistical effort. Thereby, WBE proved to complement classical surveillance programs based on individual epidemiological cases. In the long run, comprehensive WBE might be an economic way to inform traditional case-based epidemiology to adaptively adjust testing strategy and sampling density on a regional level. Furthermore, once in place, WBE provides synergies to survey prevalence and serotypes of a wider range of public health relevant pathogens, such as influenza and enterovirus ^71,72^. This will be especially relevant for countries with limited resources. In this regard, it is critical to iterate the here presented methodology to surface waters, cesspits and other modes of WW disposal in absence of a centralised sewer ^73^.

The virus conglomerates in the WW reflects the whole virus population, which differs substantially from the canonical approach of summing consensus sequences of virus isolates from individual cases. This holistic perspective on the population provides opportunities and is expected to offer additional value for WBE. It is well established in population genomics that certain applications benefit from a high sampling rate ^74^. Here, we aimed to synergise the two fields by connecting the concept of nucleotide diversity and important epidemiological parameters, such as prevalence and mobility, as a surrogate for introductions. We observed that the absolute case number is indeed imprinted into the observed nucleotide diversity. In contrast, no effect of mobility was found. Further studies are needed to clarify if this is due to a limited role of regional mobility during viral spread, or if the used data does not provide enough sensitivity for its detection. A cause for such a reduced sensitivity might be found in the pronounced effect of the selective sweep caused by the displacement by the Alpha variant, which led to a reduction of the overall nucleotide diversity. Remarkably, no similar reduction could be observed after the takeover of the Delta variant. It is important to highlight that both displacements, by Alpha and Delta, respectively, as the dominant circulating variants differed in many aspects. For example, Alpha was introduced during winter amidst a fading seasonal wave while Delta emerged during a low-incidence period in early summer, after widespread rollout of vaccines. Both replacements were accompanied by an increase in case prevalence. Interestingly, Alpha took a month to be spread across Austria and even three months to gain dominance, while Delta replaced Alpha within a month. Interestingly, during the dominance of Delta, the nucleotide diversity rose in most of the WWTP. In contrast, the nucleotide diversity remained stable during the dominance of Alpha. It is tempting to account this, beside increased population size, to a higher plasticity of Delta in contrast to Alpha. Distinctions between the phylogeny of Delta and other variants of concern are already described ^75^. The dynamic differences of Alpha and Delta emergence may hold the key for a better understanding and anticipation of future shifts in the circulating virus population.

The field of WBE, which has come a long way since its beginnings using monkeys as biosensors for poliovirus detection ^76^, received a strong impetus driven by the needs of the current pandemic management ^23,70^. Data analysis, interpretation and integration with classical epidemiological data of infectious diseases remains a formidable challenge that will require concerted interdisciplinary efforts of the scientific community, supported by water engineers, mathematicians, chemists, molecular biologists, virologists, medical scientist and computational biologists. This national-scale study demonstrates the usefulness of sequencing-based WW surveillance for the current SARS-CoV-2 pandemic and its future impact on improved global surveillance of other infectious diseases.

## Data Availability

All data produced in the present study are available upon reasonable request to the authors.

## Data and software availability

All raw sequencing data used during this study, with additional sequencing data produced for samples drawn from Austrian WWTP before December 2020, are available under the following ENA accession number: PRJEB48985. The source code for the developed and applied software for variant quantification (*VaQuERo*) and mutation deconvolution (*DeViVa*) is available on GitHub (https://github.com/fabou-uobaf/VaQuERo and https://github.com/SebH87/DeViVa).

## Acknowledgement

This project was funded in part by the Austrian Agency for Health and Food Safety (AGES), the Vienna science and Technology Fund (WWTF) as part of the WWTF COVID-19 Rapid Response Funding 2020 (A.B.), the Austrian Science Fund (FWF1212P) and the Austrian Academy of Sciences. Access to WWTP and sample logistics was possible through the Coron-A Project, which was supported and funded by the Austrian Federal Ministry of Education, Science and Research, the Austrian Federal Ministry of Agriculture, Regions and Tourism, the Provincial Governments of Lower Austria, Upper Austria, Salzburg, Carinthia, Tyrol, Styria, Burgenland, Vorarlberg and the Austrian Association of Cities and Towns. Additional samples and background information were provided by the covid-19 wastewater surveillance programs of the federal states Carinthia, Vorarlberg and Vienna. Sequencing was performed at the biomedical sequencing facility (https://www.biomedical-sequencing.org/).

## Author contributions

F.Am., R.M., L.E., S.H., L.R., M.Z., P.T., N.P., D.S., C.B. analysed data. R.M., B.A., A.S., M.Z., M.B., G.H., P.T., M.T., T.P., M.Se., J.L., Z.K., B.D., M.St., H.N., C.S., G.V., G.W., A.W., K.S., A.M., E.R., F.Al., H.O., N.K., H.I. provided processed wastewater samples, generated SARS-CoV-2 genome sequences and/or provided data. F.Am., R.M. and A.B. wrote the manuscript.

## Methods

### Analysis of WW parameters

COD, total nitrogen (tN) and ammonium nitrogen (NH_4_-N) were analysed on site by respective WWTP operators using cuvette-tests. Deposition and daily WW flow were also recorded on site.

### Sample processing

Sample drawing, RNA extraction and RT PCR-based virus quantification was performed by the members of the Coron-A consortium (www.coron-A.at), at three different laboratories (TU Wien, Medical University Innsbruck, University Innsbruck). The protocols for WW sample pre-treatment and PEG-NaCl-based precipitation of the virus fraction were based on the methods described by Ye et al. and Wu et al. ^77,78^. In details, the implemented protocols varied in each of the executing laboratory. Generally, all workflows followed a common procedure. 24 h volume equivalent mixed samples were stored at 4°C until processing. Prior to PEG-precipitation based concentration, large particles were removed. Nucleic acid extracted from pellet after concentration were analysed via one-step RT-qPCR for presence of the N-gen of SARS-CoV-2 and passed on for sequencing library preparation. Details per laboratory as follows. *University Innsbruck*: Larger particles were removed to decrease the amount of non-viral RNA and PCR-inhibitors. For this purpose, 40–70 ml of WW were transferred to centrifugation tubes and centrifuged at 4,500 g for 30 min at 4°C. To precipitate viral fragments, the resulting supernatant was immediately transferred into a fresh tube containing 10 % w/v polyethylene glycol (PEG) 8000 (CarlRoth, Karlsruhe, Germany) and 2.25 % w/v NaCl. The Reax2™ head over-head shaker (Heidolph, Schwabach, Germany) was used until both additives were dissolved. Subsequently, the samples were centrifuged at 12,000 g for 99 min at 4°C to obtain a pellet containing the viral fragments. The supernatant was removed by decantation and an additional centrifugation at 12,000 g for 5 min to remove remaining fluids with a pipette. Following the precipitation of the viral fragments, samples were resuspended in 800 μl lysis-buffer (Monarch™ total RNA Miniprep Kit, NEB, Ipswich, USA) and purified according to manufacturer protocol of the Monarch(tm) total RNA Miniprep Kit with non-enzymatic gDNA-removal. RNA was eluted in 40 μl RNase free water. RNA concentrations of the templates were quantified via Nanodrop and extracts with RNA concentrations above 200 ng/μl were diluted as needed. RNA copy numbers were determined using the N1 primers/probe according to the CDC-protocol ^79^ targeting the nucleocapsid-gene of SARS-CoV-2. RT-qPCR reactions contained per 20 μl: 10 μl Luna Universal Probe One-Step Reaction Mix (2X) from NEB, 1 μl Luna WarmStart® RT Enzyme Mix (20X) from NEB, 0.8 μl primer (final concentration 0.4 μM), 0.4 μl probe (final concentration 0.2 μM), 2 μl PCR grade water, and 5 μl template. Analyses were conducted on a RotorGene cycler (Qiagen, Hilden, Germany). After an initial reverse transcription at 55°C for 10 min, followed by 95°C for 1 min of denaturation, 45 cycles of 95°C for 10 sec and 60°C for 40 sec were performed. To calculate copy numbers, a plasmid standard containing the N-gene of SARS-CoV-2 (2019-nCoV_N_Positive Control, IDT, Leuven, Belgium) was used. ^80^

#### Medical University Innsbruck

Untreated sewage was collected with a 24 h volume-equivalent sampler, cooled to 4°C and shipped under cooled conditions to the processing laboratory. Wastewater samples were processed within 24 hours of arrival. They were stored at 4°C until analysis. In a first step, 250 ml aliquots of the WW samples were cleared by centrifugation at 4,500 x g for 30 min. In 225 ml of the resulting supernatant, 22.5 g of PEG 8000 and 5.1 g of NaCl (both Sigma-Aldrich, St. Louis, MO) were dissolved by careful swirling. These solutions were allowed to sit at 4°C for 15 min before they were spun at 12,000 g for 50 min to pellet the precipitated virus fraction. After carefully discarding the supernatant, the pellets were re-centrifuged at 12,000 g for 5 min and the residual liquid was removed with a pipette. Finally, the pellets were resuspended in 700 µl of buffer MTL (Qiagen, Hilden, Germany).

Four-hundred microliters of pellet suspension were supplemented with 60 µl of a solution comprising 60 ng carrier RNA per microliter AVL buffer (both Qiagen). Subsequently, samples were subjected to automated RNA extraction on a Biorobot EZ1 Advanced XL instrument (Qiagen) by using the EZ1 Virus Mini kit (v2.0, Qiagen) according to the manufacturer’s recommendations. Total RNA was eluted in 60 µl AVE buffer (Qiagen) and immediately used in quantitative reverse transcription real-time polymerase chain reaction (RT-qPCR) experiments targeting the N1 gene. For subsequent sequencing, 45 µl of the extracts were further purified with the Monarch(tm) total RNA Miniprep Kit (NEB, Ipswich, USA) according to the manufacturer’s protocol. Total RNA was eluted in 45 µl water.

#### TU Wien

Thoroughly mixed 45 ml aliquots from each sample were centrifuged at 4,500 g for 30 min at 4°C (the centrifuge was previously cooled down to 4°C). 40 ml of the supernatant were further added to fresh 50 ml falcon tubes containing 100 g/L PEG (Sigma Aldrich) 8000 and 22.5 g/L NaCl (Sigma Aldrich), followed by centrifugation at 12,000 g for 1h 30 min at 4°C. The supernatant was carefully removed, and an additional centrifugation step was carried out in order to remove the remaining liquid (12,000 g, 5 min, 4°C). The pellet was resuspended in 400 µl molecular biology grade water (Sigma Aldrich) and 400 µl CTAB (Hexadecyltrimethylammonium bromide; Promega) buffer. Prior to the RNA extraction, 40 µl of Proteinase K (Promega) were added to each tube and inverted carefully in order to obtain a homogenous suspension. The suspension was further transferred to a tube containing disruptor beads and run on FastPrep24 (MP Biomedicals™) machine for 40 s at 6 m/s. Subsequently, 400 µl of each sample were used for further RNA extraction. The RNA extraction was performed on Maxwell^®^ RSC AS4500 (Promega) device using Viral RNA/DNA Concentration and Extraction Kit from Wastewater (Promega). The SARS-CoV-2 viral RNA was eluted in 100 µl TE buffer.

### RNA library prep and sequencing

For amplicon cleanup AMPure XP beads (Beckman Coulter) were used at a 1:1 ratio and amplicon concentrations and size distributions were checked with the Qubit Fluorometric Quantitation (Life Technologies) and the 2100 Bioanalyzer system (Agilent), respectively. Amplicon concentrations were normalized, and sequencing libraries created with the NEBNext Ultra II DNA Library Prep Kit for Illumina (NEB) according to the manufacturer’s instructions. As before library concentrations and size distribution were assessed again, the pools mixed at equimolar concentrations and sequencing carried out on the NovaSeq 6000 platform (Illumina) using an SP flow cell with a read length of 2×250 bp in paired-end mode.

### Mutation calling

After demultiplexing, adapter sequences were trimmed with BBDUK, and the overlapping regions of paired reads were corrected with BBMERGE from the BBTools suite ^81^. BWA-MEM ^82^ (v 0.7.17) was then used to map read pairs to the combined GRCh38 and SARS-CoV-2 genomes (RefSeq: NC_045512.2) with a minimal seed length of 17. Only reads mapping uniquely and properly paired to the SARS-CoV-2 genome were kept and primer sequences were masked with iVar ^83^. LoFreq ^84^ (v 2.1.2) was used for low frequency variant calling, after first using its Viterbi method to realign reads around insertions and deletions (InDels) and adding InDel qualities. Variants were then filtered with LoFreq and Bcftools ^85^ (v 1.9) only considering variants with a minimum coverage of 75 reads, a minimum Phred scaled calling quality value of 90 and InDels with a HRUN value of at less than 4. Variant annotation was performed with SnpEff ^86^ (v 4.3) and SnpSift ^87^ (v 4.3).

### Virus variant quantification, including marker mutation definition

Multiple genome alignments of 1.9 million global SARS-CoV-2 genomes provided by the global initiative on sharing avian influenza data (GISAID database, retrieved July 5^th^, 2021) ^58,59^, and the associated pangolin lineage assignments, were used to extracted all nucleotide variants compared to the reference genome. For each pangolin lineage, all mutations that could be observed in at least 85% of the analysed genomes were defined as sensitive marker mutations for the respective lineage. To reduce complexity, the scope was focused on all lineages which were detected in Austria (at least ten sequenced genomes according to GISAID) and all variants of concern/interest and variants under monitoring as defined by the European Centre for Disease Prevention and Control (ECDC; https://www.ecdc.europa.eu/en/covid-19/variants-concern; accessed on July 7^th^, 2021). A total of 36 different regionally relevant variants were included (**Extended Data Fig. 4**). Deduced marker mutation, together with inferred mutation frequencies, were further utilized to quantify all respective variants of relevance per WWTP in the available time course data according to the following scheme: (i) remove all samples with more than 60% of the genomic position covered with less than 10 reads; (ii) filter marker mutations with observed frequency >0.02 and >75 supportive reads; (iii) foreach timepoint, denote all variants with at least three unique mutations as detected; (iv) add mutation frequencies from samples from the preceding 8 days; (v) filter frequencies of marker mutation of detected variants; (vi) transform ^88^ remaining mutation frequencies to avoid zeros and ones by *f*′ = (*f*(*d* − 1) + 0.5)/*d*, whereas d denotes the sequencing depth at the respective locus, f and f’ denote the observed and the transformed mutation frequency, respectively; (vii) infer expected frequency per variant applying a regression model y = Xβ, whereas y represents the inferred variant frequencies, b the mutation frequencies and X represents the *n* x *m* design matrix assigning the considered *n* mutations to one or more of the *m* variants. To this end the package gamlss ^89^ (generalized additive model for location scale and shape) is used with a SIMPLEX ^90^ linker function and weights *w* per mutation i 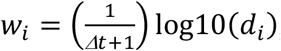, with the time difference to the current sample in focus in days Δt and the sequencing depth d_i_ for at mutation position i. If the SIMPLEX function does not converge, resort to the Beta function. If sum of expected values exceeds 1, scale down proportionally. An implementation of the described software is available on GitHub (https://github.com/fabou-uobaf/VaQuERo). Spatiotemporal reconstruction was performed from the deduced variant frequencies across Austria using the Multilevel B-Spline Approximation ^91^ using the mba.surf function implemented in the R package MBA with the following non-default parameters: no.X = no.Y = 30, m = range of longitude [°] divided by range of date [d], and extend set to TRUE.

### Deduction of a variant specific reproduction number from WW

SARS-CoV-2 load was normalized to the ammonium nitrogen (NH_4_-N) load as population size marker, assuming a NH_4_-N load per capita of 8 g per day ^64^. The normalization results in the viral load parameter λ, representing viral copies per population equivalent per day, which is proportional to the absolute number of infected individuals ^92^. In combination with the relative proportions from the sequencing based variant quantification (derived from the *VaQuERo*), the absolute quantity λ_*v*_ from a certain variant *v* was estimated. Time series at a daily base, necessary for the following R_ww_ calculation, were interpolated by LOESS smoothing (smoothing factor 0.25). Subsequently, R_ww_ was calculate using the EpiEstim Package ^65^ (https://github.com/mrc-ide/EpiEstim) based on λ _*v*_, applying a fixed serial interval of 3.37 d ± 1.83 (see ref. ^93^). The R_ww_ was separately calculated for the Alpha variant 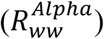 and the sum of all other pre-Alpha variants 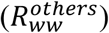. The ratio 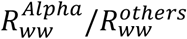, as a measure for relative fitness disparity, was analysed for the transition period January to June 2021 during which Alpha gained dominance in Austria.

### Evaluation by epidemiological records

The national Covid-19 surveillance system, maintained by AGES, consist of all confirmed SARS-CoV-2 cases since the beginning of the pandemic. In the period of examination between Jan 1^st^ and Sep 15^th^, 2021, on average 70.8% of all identified SARS-CoV-2 cases in all towns connected to any examined WWT were tested for SARS-CoV-2 mutations and variants. Sequencing based tests were performed by partial sequencing of the spike gene ^94^, Sanger sequencing or WGS. However, most cases were analysed using PCRs for specific mutations, performed in the responsible diagnosis laboratory and reported to AGES. Based on the set of reported mutations a case was classified as a particular variant of concern or as the wild type, accordingly. Definition of indicator mutations were adapted during the pandemic. Individual cases are associated to their place of residence. Data were extracted from the surveillance system for all towns connected to any examined WWTP and aggregated for all towns within the same catchment area and from the same calendar week, to deduce absolute case counts and relative frequency per variant per catchment area per week. For plotting purpose, and for direct comparison with WW derived data, both epidemiological and WW data was shifted to Wednesday of that respective week.

### Mutation Deconvolution

For mutation deconvolution, a hierarchical, unsupervised two-step clustering approach was used. First, a silhouette-analysis collected mutations with similar pattern of relative abundances within a specific set of samples. The method is implemented in a software tool named *DeViVa* (Deconvolution of Virus Variants), which was written in python (version 3.8) and which can be obtained from GitHub (https://github.com/SebH87/DeViVa). The first clustering step used Ward’s method based on Euclidian distance in order to shed spurious mutations. To guarantee consistent results, only mutations with a relative abundance >5% were accepted. The relative abundance of the remaining mutations was transformed (centered Log-ratio transformation; 0 was replaced by 10^−7^) and subjected to a second hierarchical clustering step. Therefore, distances between mutations were calculated by applying the complete-linkage algorithm based on square Euclidian distance. To identify distinct mutation constellations a silhouette analysis was performed. Thereby, a silhouette coefficient is calculated for each clustering produced by a critical distance threshold between 2 and 20. The clustering exhibiting the highest silhouette coefficient is considered optimal and is further used. Finally, each cluster is variant-typed by introducing its mutations into the reference-genome of SARS-CoV-2 and analysed by the Pangolin-lineage tool ^54^.

### Nucleotide diversity

The loFreq filtered variant call format files of all WWTP for which more than 18 timepoints were available with at least 2/3 of the genome covered with more than ten reads, were used to extract variants with an allele frequency above 1%. Multiallelic SNPs were joined using Bcftools ^85^ (v1.12). SNPgenie ^95^ (v2019.10.31) was used to calculate genome wide nucleotide diversity π. For samples with more than 18 timepoints, the data was integrated with abundances of variant of concern as deduced by *VaQuERo*. Significance of differences in πbetween different periods was tested using a Mann-Whitney U test between π-values from ± 4 weeks around observed introduction of Alpha and from ± 4 weeks around maximal observed dissemination of Alpha.

### Mobility data

Statistics on registered accommodation in the hospitality industry (Beherbergungsstatistik 2019/20, https://www.statistik.at/atlas/?mapid=them_tourismus_winter_uebernachtungen), and the statistic on registered commuting dynamics (Registerzählung 2011, Abgestimmte Erwerbsstatistik, https://www.statistik.at/atlas/?mapid=them_bevoelkerung_pendler) were gathered from publicly available databases. Of note, both registers do not represent the same period, and moreover, it is incalculable how social adaptation (work from home and circumvention of hospitality restrictions) has affected the registered behaviour. Therefore, mobility data based on the mobile communications network records from one undisclosed mobile provider were furthermore used. A movement from region A to B in the origin destination matrix was measured if a mobile phone was first logged in to A for at least 15 minutes and afterwards logged in to B for at least 15 minutes. Detailed information about the origin destination matrix calculation process have been described previously ^96^. This movement data was used to deduce the proportion of population who at least once per day leaves the catchment area in the direction of another area or even another federal state. Therefore, the number of movements leaving the districts which are assigned to the WWTP were counted for each catchment. The retrieved statistics were normalized to the number of people connected to the sewage system and log10 transformed.

## Extended Data

**Extended Data Fig. 1.**
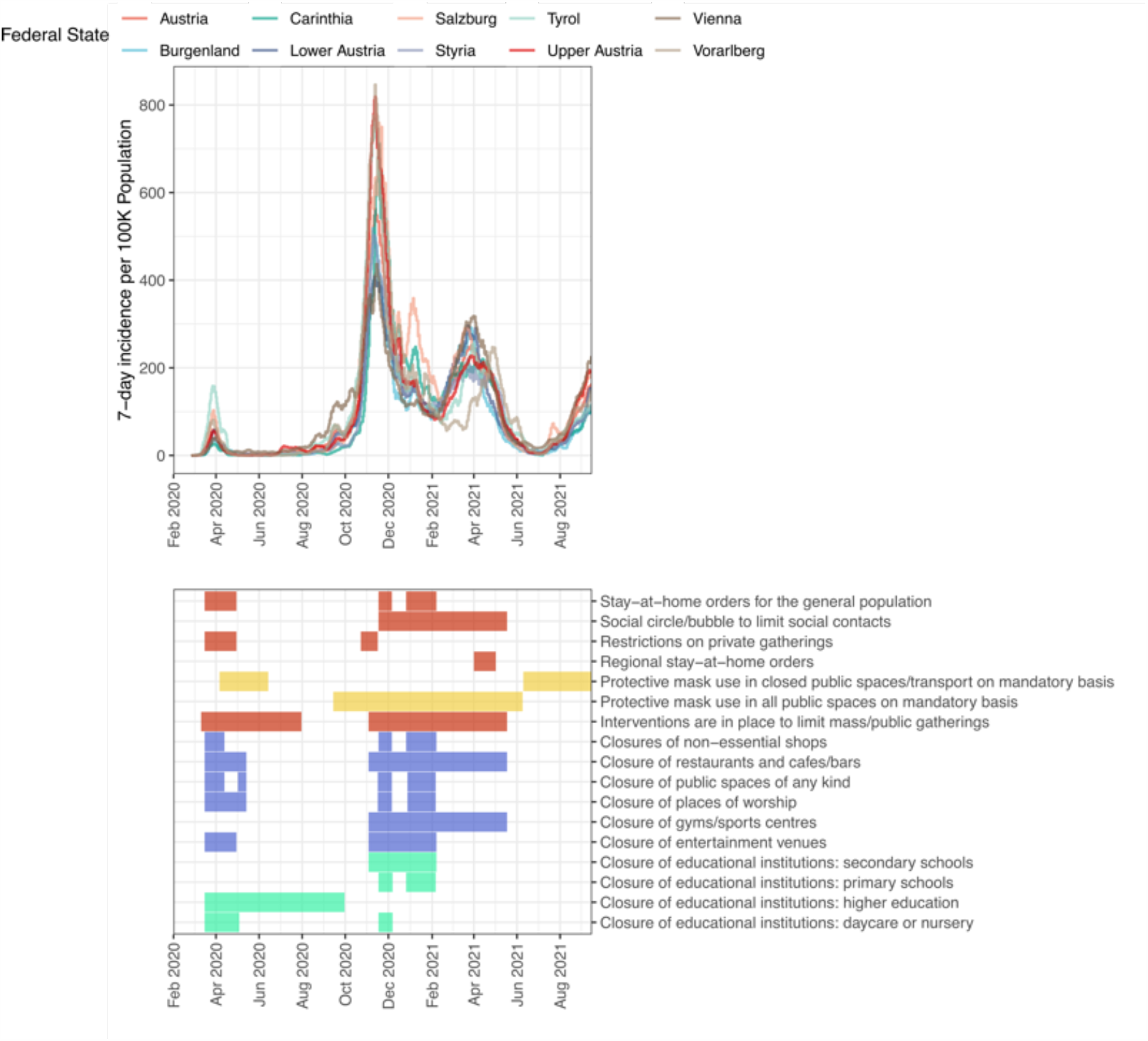
Course of the COVID-19 pandemic in Austria. Upper panel depicts the average daily cases per 100K population in last 7 days for the course of the COVID-19 pandemic for each Austrian province and for all of Austria (https://covid19-dashboard.ages.at). The lower panel indicates the non-pharmaceutical interventions in the same time period (adapted from https://www.ecdc.europa.eu/sites/default/files/documents/response_graphs_data_2021-10-21.csv).

**Extended Data Fig. 2.**
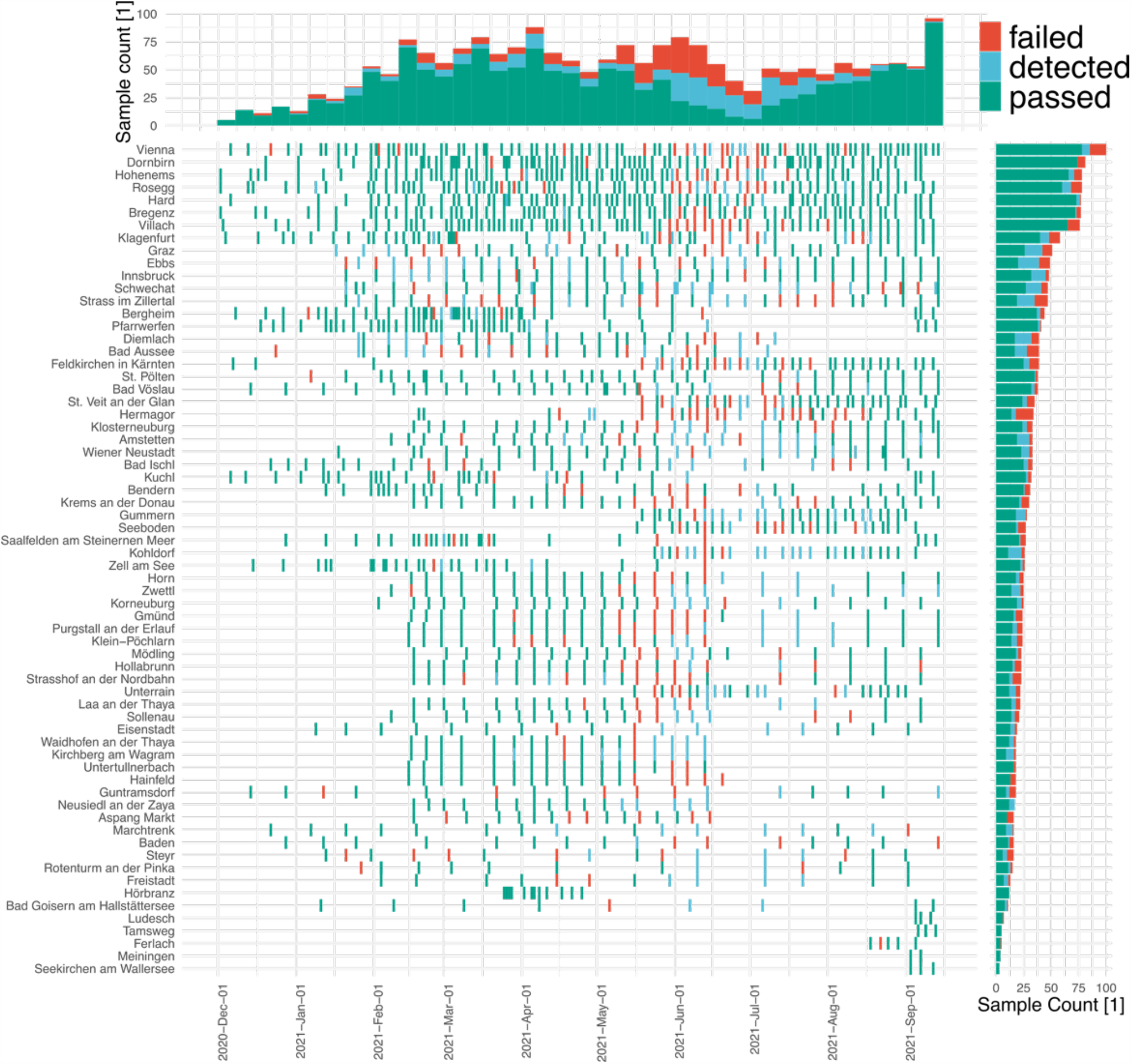
Data collection. Graphic representation of all sequenced samples from WWTP with more than three timepoints. Sample date and location, and sequencing success is indicated. Samples are classified into *passed* and *detected* if 40% and 5% of the genome, respectively, is covered with more than 10 reads. A coverage of more than 5% of the genome was considered as robust detection of the virus, since at least three independent amplicons responded, but too little information is provided to reliably characterize viral variants. Therefore, detected samples were not used further. If less than 5% of the genome was covered the sequencing of the respective samples was categorized as *failed*. 70.7% of all samples were classified as *passed*. Another 14.6% and 14.7% were classified as *detected* and *failed*, respectively.

**Extended Data Fig. 3.**
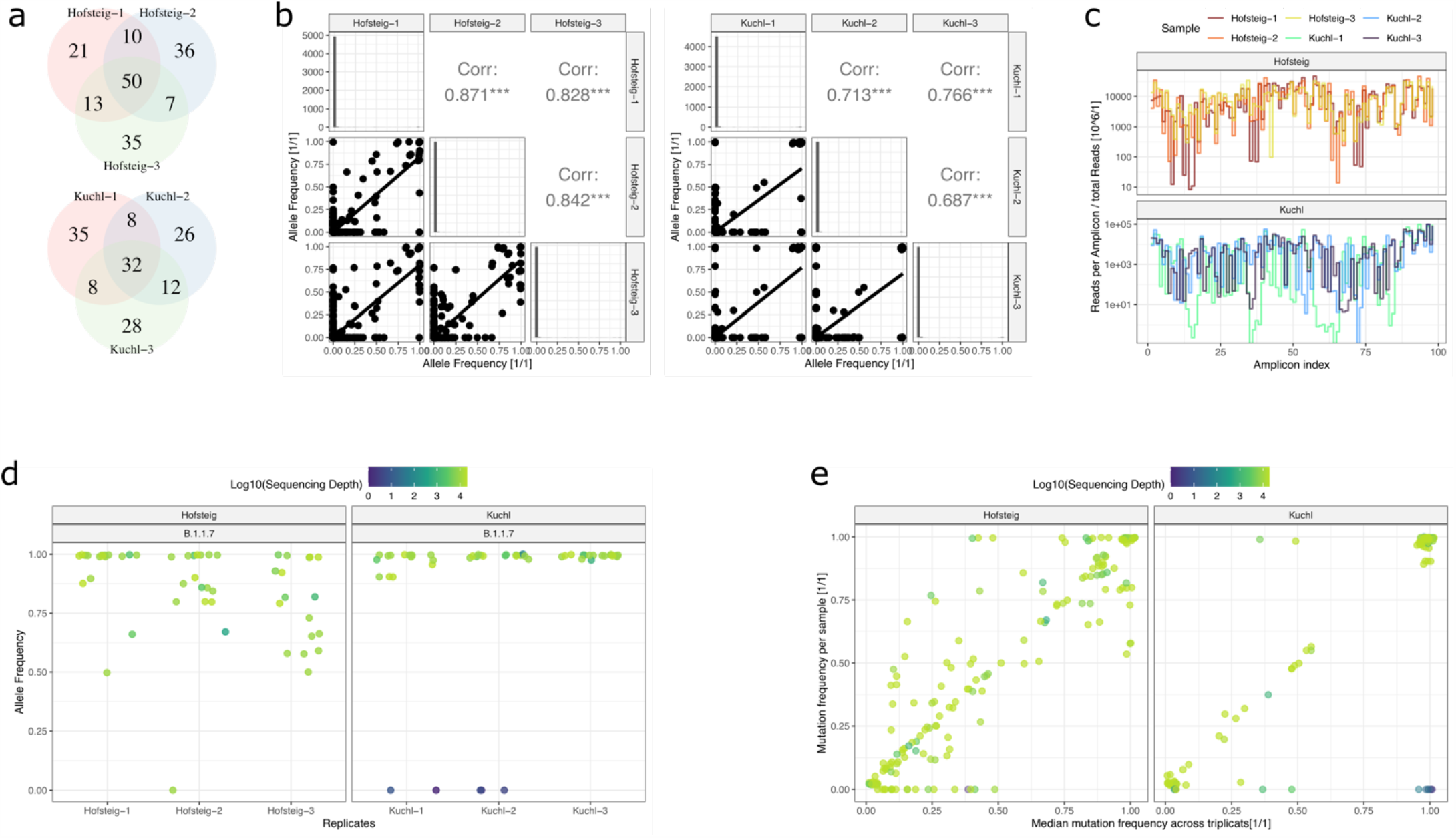
Reproducibility analysis. Analysis of triplicate sample sets collected on the same day at two different WWTP and processed independently. **a**, Reproducibility analysis for detection of mutations. Venn diagram over the sets of mutations observed with a frequency greater zero in an individual sample for all mutations called in any of the triplicates with an allele frequency greater 2%. **b**, Analysis of triplicate samples with respect to the qualitative reproducibility of observed allele frequencies. Correlation between allele frequencies is modest with correlation factors between 0.69 and 0.87. Generally, mutation dropout in one sample contributes heavily to a reduced correlation factor. **c**, Reproducibility of normalized read-coverage per amplicon between the triplicates. **d**, deduced allele frequencies of marker mutations of the Alpha (B.1.1.7) variant (dominating variant according to WW and epidemiological data at the corresponding time) in the triplicates. Point colour indicates read depth at the respective loci. **e**, scatter plot depicting the median mutation frequency and the individual mutation frequencies for each detected mutation (with median frequency greater than 2%) in the two replication sets. Samples at the baseline (y = 0) represent complete dropouts in one replicate. Point colour indicates read depth at the respective loci. As in panel d, some mutation with expected presence according to the replicates are absent even though the position is covered by a significant number of reads.

**Extended Data Fig. 4.**
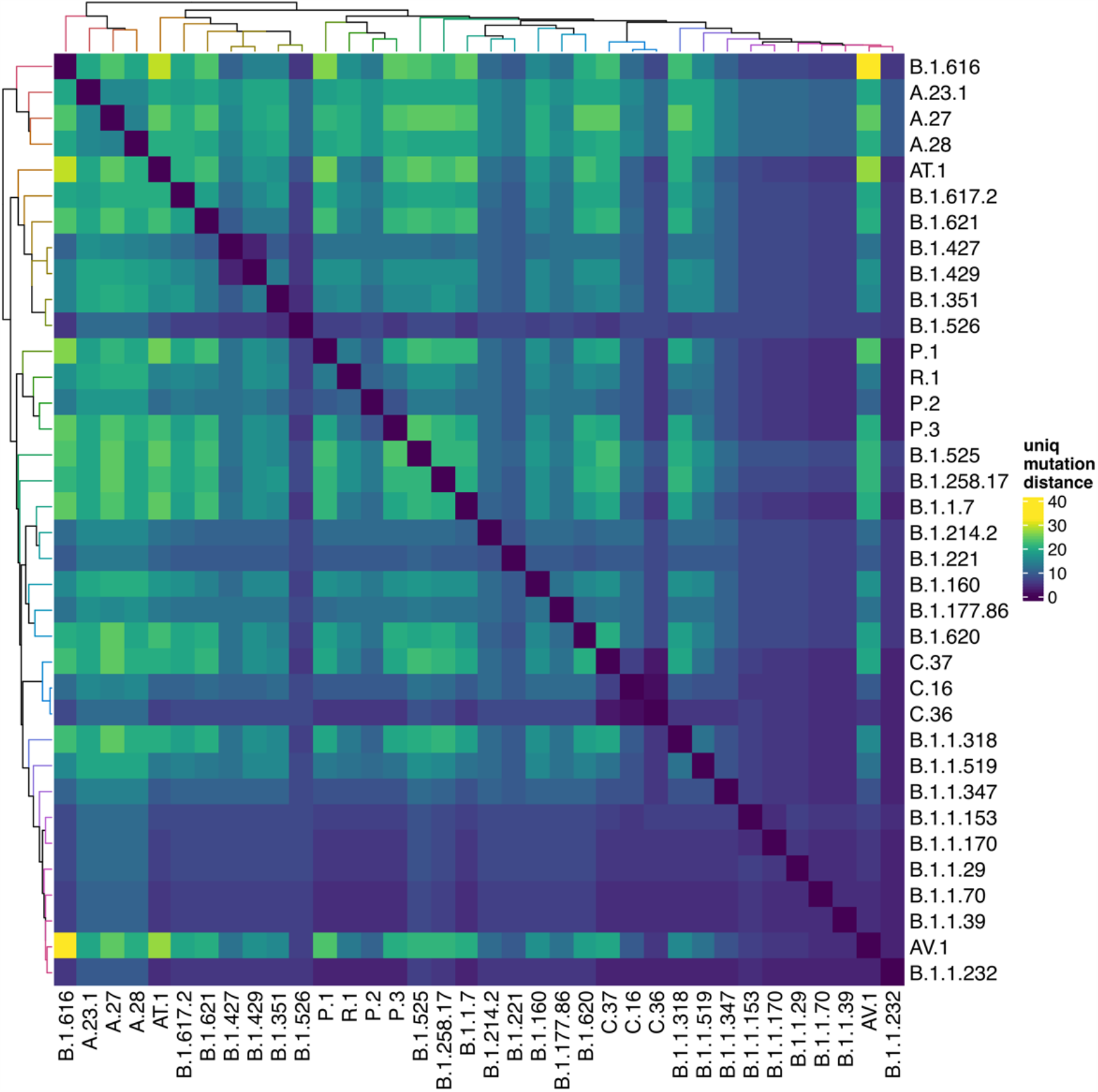
Variants of relevance. Graphic representation of all variants of relevance whose abundance in Austrian WW samples were examined. Variants which were defined by ECDC to be variants of concern, variants of interest or variants under monitoring were included. Additionally, all variants which were observed in more than ten Austrian patient samples were added. For each variant sensitive marker mutations were defined based on their occurrence (in >85% of all samples of the respective lineage) and specificity (unique to one lineage). The heatmap depicts the number of unique (specific) marker mutations between each pair of variants of relevance.

**Extended Data Fig. 5.**
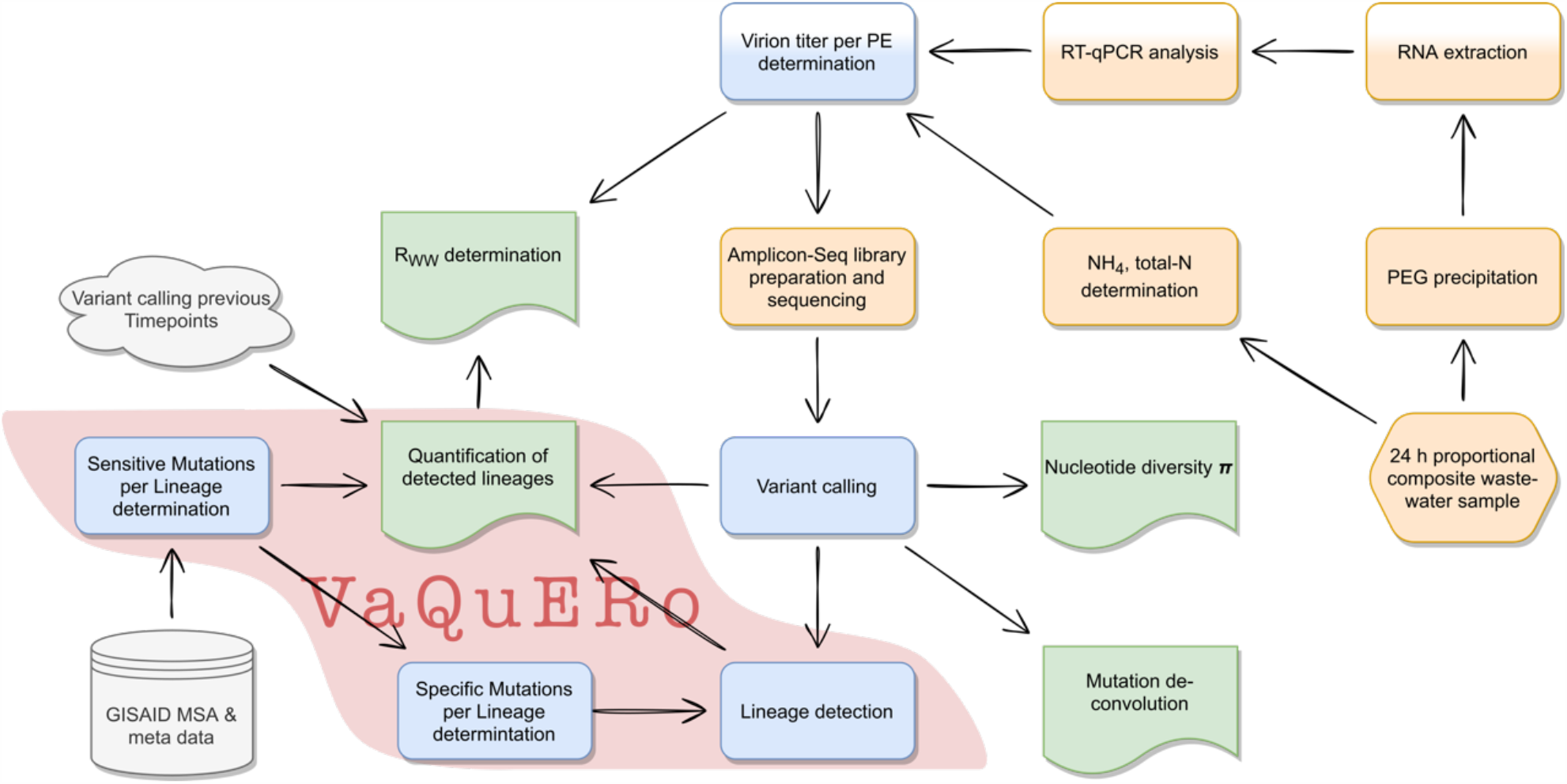
Analysis workflow. Illustration of the main analysis workflow steps from the WW sampling point, the wet lab procedures (orange panels), via the bioinformatics analysis steps (blue panels) to the analysis results (green panels). Major read outs of the analysis are nucleotide diversity π, cluster of correlated mutation constellations, variant quantification, and determination of reproduction number in the catchment area. Analysis steps implemented in the software tool *VaQuERo* are highlighted by a red area.

**Extended Data Fig. 6.**
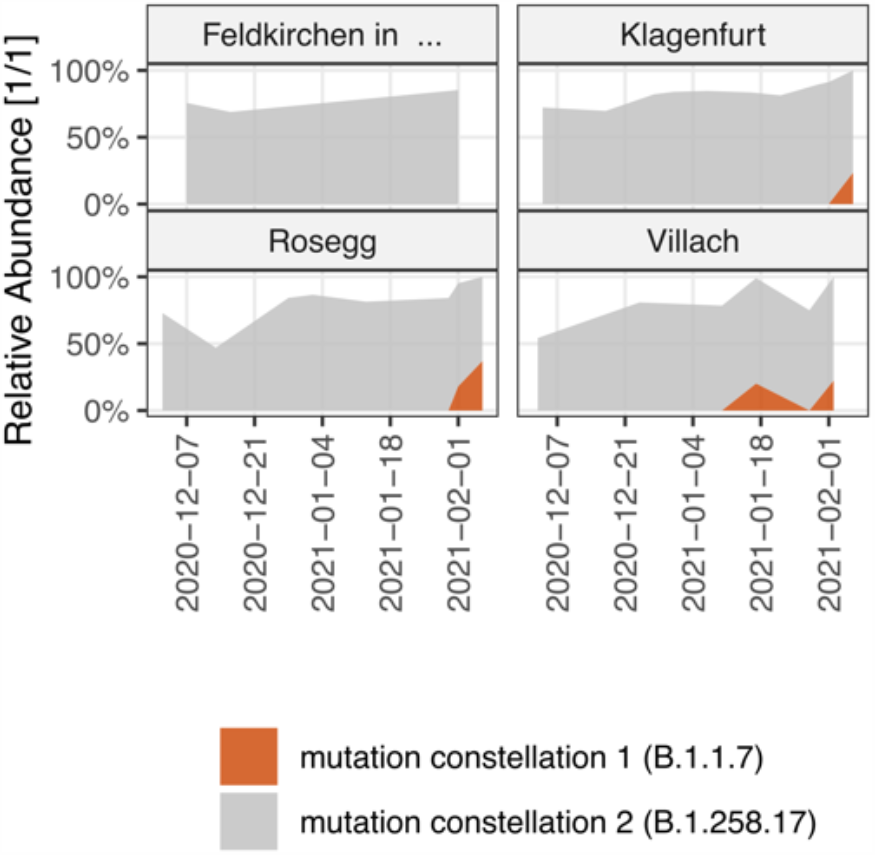
Mutation constellation deconvolution. Mutations which are grouped into related mutation constellation based on their mutation frequencies, as depicted in Fig. 3a, are used as input for *VaQuERo*, instead of the normal GISAID trained variant defining mutations. The graphic illustrates the geographic distribution and the dynamics of the de novo inferred mutation constellation enriched in Alpha (B.1.1.7) associated mutations.

**Extended Data Fig. 7.**
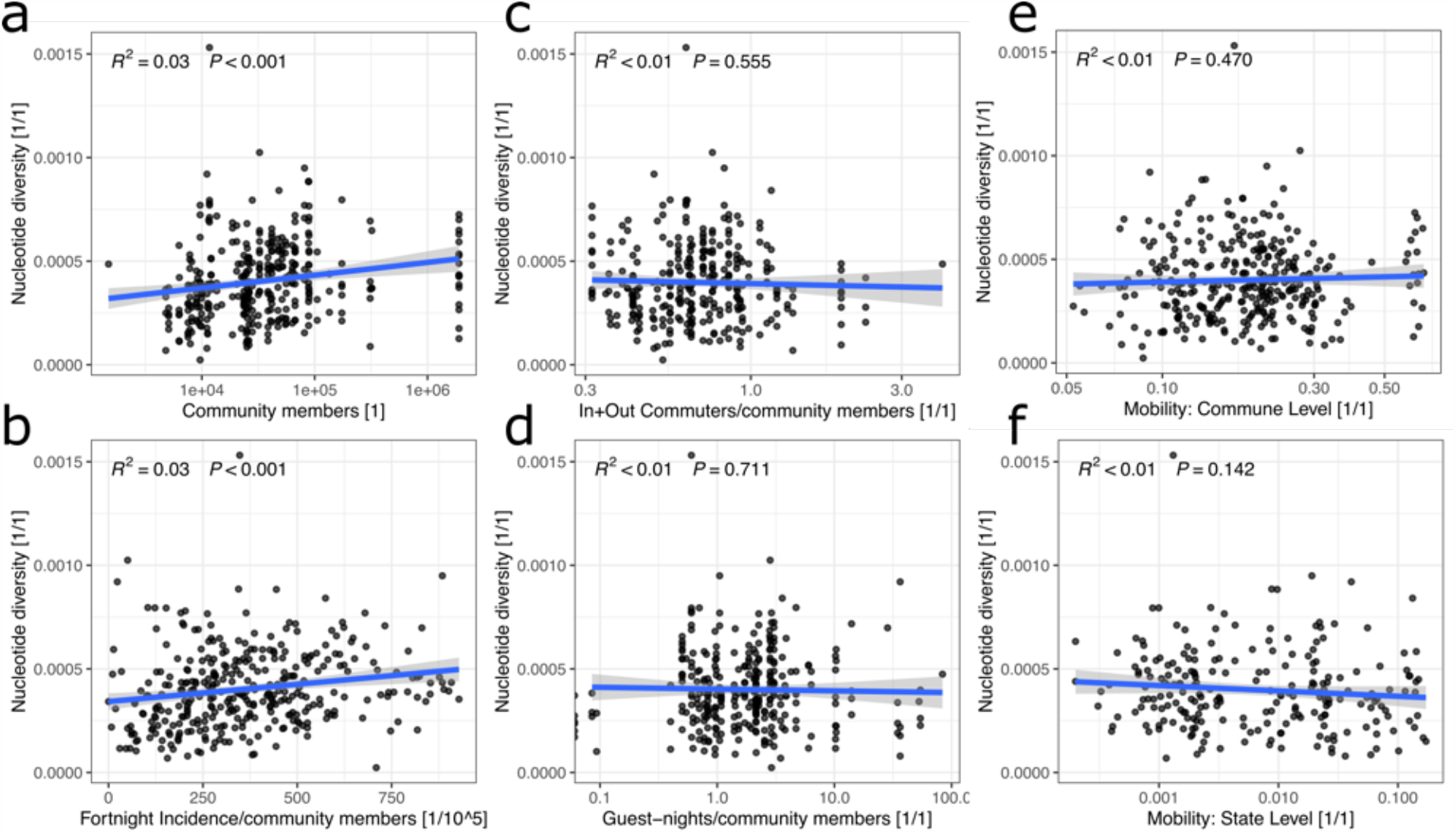
Virus population diversity and mobility. Correlation analysis between nucleotide diversity measure π of all samples with quantified Alpha variant above 95% and external data, describing (from left to right) the number of people in the catchment area **a**, people connected to the WWTP in the catchment area. **b**, the 14-day incidence rate per 100,000 people **c**, the proportion of in and out commuters (as officially registered) to the total number of people in the catchment area **d**, the number of registered over-night days in the hospitality industry in the catchment area **e**, the proportion of population leaving the commune **f**, or the federal state, as observed by mobile communications network records.

**Extended Data Fig. 8.**
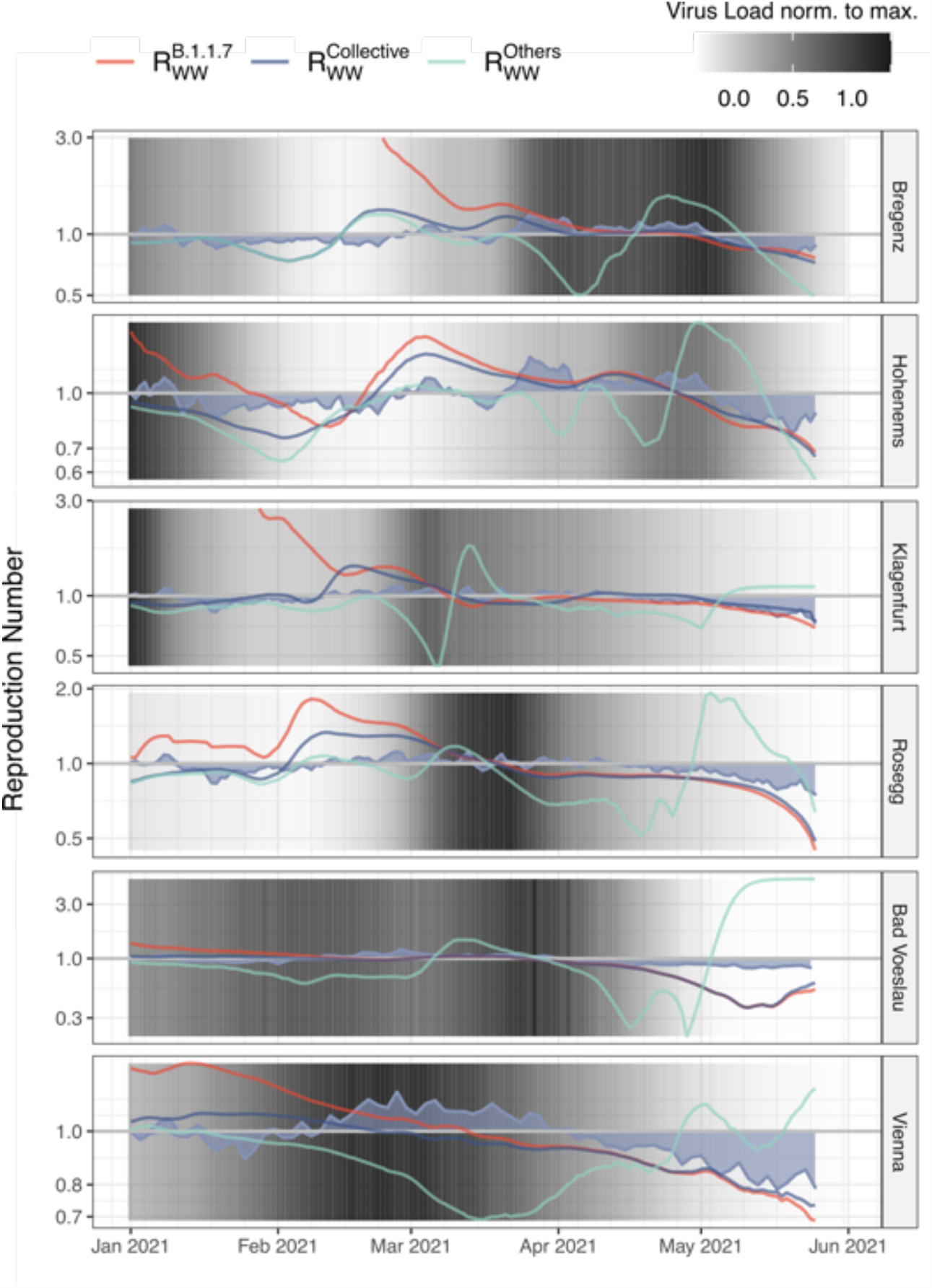
Reproduction number R_ww_ deduced from WW. Selected WWTP with the deduced variant-specific reproduction number R_ww_. The gauged virus load (cp/PE) as observed in the WW is indicated with background shades. The blue area indicates the estimated R_eff_ per respective federal state as deduced from epidemiological case monitoring.

## References

1. Nicola, M. et al. The socio-economic implications of the coronavirus pandemic (COVID-19): A review. International Journal of Surgery vol. 78 185–193 (2020).

2. Josephson, A., Kilic, T. & Michler, J. D. Socioeconomic impacts of COVID-19 in low-income countries. Nature Human Behaviour 5, 557–565 (2021).

3. Case, J. B., Winkler, E. S., Errico, J. M. & Diamond, M. S. On the road to ending the COVID-19 pandemic: Are we there yet? Virology 557, 70–85 (2021).

4. Krammer, F. SARS-CoV-2 vaccines in development. Nature vol. 586 516–527 (2020).

5. Harvey, W. T. et al. SARS-CoV-2 variants, spike mutations and immune escape. Nature Reviews Microbiology vol. 19 409–424 (2021).

6. Callaway, E. Heavily mutated coronavirus variant puts scientists on alert. Nature (2021).

7. Martin, D. P. et al. The emergence and ongoing convergent evolution of the N501Y lineages coincides with a major global shift in the SARS-CoV-2 selective landscape. medRxiv : the preprint server for health sciences (2021) doi:10.1101/2021.02.23.21252268.

8. Valesano, A. L. et al. Temporal dynamics of SARS-CoV-2 mutation accumulation within and across infected hosts. PLoS Pathogens 17, (2021).

9. Truong, T. T. et al. Increased viral variants in children and young adults with impaired humoral immunity and persistent SARS-CoV-2 infection: A consecutive case series. EBioMedicine 67, (2021).

10. Luo, R., Delaunay-Moisan, A., Timmis, K. & Danchin, A. SARS-CoV-2 biology and variants: anticipation of viral evolution and what needs to be done. Environmental Microbiology 23, 2339– 2363 (2021).

11. Chandler, J. C. et al. SARS-CoV-2 exposure in wild white-tailed deer (Odocoileus virginianus). doi:10.1101/2021.07.29.454326.

12. Lucas, C. et al. Impact of circulating SARS-CoV-2 variants on mRNA vaccine-induced immunity. Nature (2021) doi:10.1038/s41586-021-04085-y.

13. Furuse, Y. Genomic sequencing effort for SARS-CoV-2 by country during the pandemic. International Journal of Infectious Diseases 103, 305–307 (2021).

14. The COVID-19 Genomics UK (COG-UK) consortium. An integrated national scale SARS-CoV-2 genomic surveillance network. The Lancet Microbe vol. 1 e99–e100 (2020).

15. Treibel, T. A. et al. COVID-19: PCR screening of asymptomatic health-care workers at London hospital. The Lancet 395, 1608–1610 (2020).

16. Brito, A. F. et al. Global disparities in SARS-CoV-2 genomic surveillance. medRxiv : the preprint server for health sciences (2021) doi:10.1101/2021.08.21.21262393.

17. Belman, S., Saha, S. & Beale, M. A. SARS-CoV-2 genomics as a springboard for future disease mitigation in LMICs. Nature Reviews Microbiology (2021) doi:10.1038/s41579-021-00664-y.

18. Majid, F., Omer, S. B. & Khwaja, A. I. Optimising SARS-CoV-2 pooled testing for low-resource settings. The Lancet Microbe vol. 1 e101–e102 (2020).

19. Larsen, D. A., Green, H., Collins, M. B. & Kmush, B. L. Wastewater monitoring, surveillance and epidemiology: a review of terminology for a common understanding. FEMS Microbes 2, (2021).

20. Sharma, V. K. et al. COVID-19 epidemiologic surveillance using wastewater. Environmental Chemistry Letters vol. 19 1911–1915 (2021).

21. Foladori, P. et al. SARS-CoV-2 from faeces to wastewater treatment: What do we know? A review. Science of the Total Environment 743, (2020).

22. Chen, Y. et al. The presence of SARS-CoV-2 RNA in the feces of COVID-19 patients. Journal of Medical Virology 92, 833–840 (2020).

23. Bonanno Ferraro, G. et al. A State-of-the-Art Scoping Review on SARS-CoV-2 in Sewage Focusing on the Potential of Wastewater Surveillance for the Monitoring of the COVID-19 Pandemic. Food and Environmental Virology (2021) doi:10.1007/s12560-021-09498-6.

24. Hassard, F., Lundy, L., Singer, A. C., Grimsley, J. & di Cesare, M. Innovation in wastewater near-source tracking for rapid identification of COVID-19 in schools. The Lancet Microbe vol. 2 e4–e5 (2021).

25. la Rosa, G. et al. SARS-CoV-2 has been circulating in northern Italy since December 2019: Evidence from environmental monitoring. Science of the Total Environment 750, (2021).

26. Martin, J. et al. Tracking SARS-CoV-2 in sewage: Evidence of changes in virus variant predominance during COVID-19 pandemic. Viruses 12, (2020).

27. Nemudryi, A. et al. Temporal Detection and Phylogenetic Assessment of SARS-CoV-2 in Municipal Wastewater. Cell Reports Medicine 1, (2020).

28. Wurtzer, S. et al. Monitoring the propagation of SARS CoV2 variants by tracking identified mutation in wastewater using specific RT-qPCR. medRxiv 2021.03.10.21253291 (2021) doi:10.1101/2021.03.10.21253291.

29. Agrawal, S., Orschler, L. & Lackner, S. Long-term monitoring of SARS-CoV-2 RNA in wastewater of the Frankfurt metropolitan area in Southern Germany. Scientific reports 11, 5372 (2021).

30. Peccia, J. et al. Measurement of SARS-CoV-2 RNA in wastewater tracks community infection dynamics. Nature Biotechnology 38, 1164–1167 (2020).

31. Karthikeyan, S. et al. High-Throughput Wastewater SARS-CoV-2 Detection Enables Forecasting of Community Infection Dynamics in San Diego County. mSystems 6, (2021).

32. Izquierdo-Lara, R. et al. Monitoring SARS-CoV-2 circulation and diversity through community wastewater sequencing, the netherlands and belgium. Emerging Infectious Diseases 27, 1405–1415 (2021).

33. Crits-Christoph, A. et al. Genome Sequencing of Sewage Detects Regionally Prevalent SARS-CoV-2 Variants. (2021) doi:10.1128/mBio.

34. Agrawal, S. et al. A pan-European study of SARS-CoV-2 variants in wastewater 2 under the EU Sewage Sentinel System. doi:10.1101/2021.06.11.21258756.

35. Bar-Or, I. et al. Detection of SARS-CoV-2 variants by genomic analysis of wastewater samples in Israel. Science of the Total Environment 789, (2021).

36. Fontenele, S. et al. High-throughput sequencing of SARS-CoV-2 in wastewater provides insights into 1 circulating variants 2 3 Rafaela. doi:10.1101/2021.01.22.21250320.

37. Fuqua, J. L. et al. A rapid assessment of wastewater for genomic surveillance of SARS-CoV-2 variants at sewershed scale in Louisville, KY. medRxiv : the preprint server for health sciences (2021) doi:10.1101/2021.03.18.21253604.

38. Jahn, K. et al. Detection of SARS-CoV-2 variants in Switzerland by genomic analysis of wastewater samples. doi:10.1101/2021.01.08.21249379.

39. Pechlivanis, N. et al. Detecting SARS-CoV-2 lineages and mutational load in municipal wastewater; a use-case in the metropolitan area of Thessaloniki, Greece. 1–14 doi:10.1101/2021.03.17.21252673.

40. Smyth, D. S. et al. Detection of Mutations Associated with Variants of Concern Via High Throughput Sequencing of SARS-CoV-2 Isolated from NYC Wastewater 2 3. doi:10.1101/2021.03.21.21253978.

41. Crits-Christoph, A. et al. Genome Sequencing of Sewage Detects Regionally Prevalent SARS-CoV-2 Variants. (2021) doi:10.1128/mBio.

42. Agrawal, S., Orschler, L. & Lackner, S. Long-term monitoring of SARS-CoV-2 RNA in wastewater of the Frankfurt metropolitan area in Southern Germany. Scientific reports 11, 5372 (2021).

43. Izquierdo-Lara, R. et al. Monitoring SARS-CoV-2 circulation and diversity through community wastewater sequencing, the netherlands and belgium. Emerging Infectious Diseases 27, 1405–1415 (2021).

44. la Rosa, G. et al. Rapid screening for SARS-CoV-2 variants of concern in clinical and environmental samples using nested RT-PCR assays targeting key mutations of the spike protein. Water Research 197, (2021).

45. Nemudryi, A. et al. Temporal Detection and Phylogenetic Assessment of SARS-CoV-2 in Municipal Wastewater. Cell Reports Medicine 1, (2020).

46. Prado, T. et al. Wastewater-based epidemiology as a useful tool to track SARS-CoV-2 and support public health policies at municipal level in Brazil. Water Research 191, (2021).

47. Rimoldi, S. G. et al. Presence and infectivity of SARS-CoV-2 virus in wastewaters and rivers. Science of the Total Environment 744, (2020).

48. Agrawal, S., Orschler, L. & Lackner, S. Metatranscriptomic Analysis Reveals SARS-CoV-2 Mutations in Wastewater of the Frankfurt Metropolitan Area in Southern Germany. Microbiology Resource Announcements 10, (2021).

49. Jahn, K. et al. Detection and surveillance of SARS-CoV-2 genomic variants in wastewater. doi:10.1101/2021.01.08.21249379.

50. Huisman, J. S. et al. Wastewater-based estimation of the effective reproductive number of SARS-CoV-2. doi:10.1101/2021.04.29.21255961.

51. Kreidl, P. et al. Emergence of coronavirus disease 2019 (COVID-19) in Austria. Wiener Klinische Wochenschrift 132, 645–652 (2020).

52. Popa, A. et al. Genomic epidemiology of superspreading events in Austria reveals mutational dynamics and transmission properties of SARS-CoV-2. Sci. Transl. Med vol. 12 https://www.science.org (2020).

53. Haug, N. et al. Ranking the effectiveness of worldwide COVID-19 government interventions. Nature Human Behaviour 4, 1303–1312 (2020).

54. O’Toole, Á. et al. Assignment of epidemiological lineages in an emerging pandemic using the pangolin tool. Virus Evolution 7, (2021).

55. Klikovits, J. et al. GISAID Österreich-Report Nr. 6 (https://www.ages.at/download/0/0/fbdce8f6e79df96cf599d90959e117034e60d123/fileadmin/AGES2015/Wissen-Aktuell/COVID19/GISAID-Report_Nummer-6.pdf). (2021).

56. Özkan, E. et al. High-throughput Mutational Surveillance of the SARS-CoV-2 Spike Gene. doi:10.1101/2021.07.22.21259587.

57. Hasell, J. et al. A cross-country database of COVID-19 testing. Scientific Data 7, (2020).

58. Elbe, S. & Buckland-Merrett, G. Data, disease and diplomacy: GISAID’s innovative contribution to global health. Global Challenges 1, 33–46 (2017).

59. Shu, Y. & McCauley, J. GISAID: Global initiative on sharing all influenza data – from vision to reality. Eurosurveillance vol. 22 (2017).

60. Cragg, J. G. Some Statistical Models for Limited Dependent Variables with Application to the Demand for Durable Goods. vol. 39 (1971).

61. van Poelvoorde, L. A. E. et al. Strategy and Performance Evaluation of Low-Frequency Variant Calling for SARS-CoV-2 Using Targeted Deep Illumina Sequencing. Frontiers in Microbiology 12, (2021).

62. Itokawa, K., Sekizuka, T., Hashino, M., Tanaka, R. & Kuroda, M. Disentangling primer interactions improves SARS-CoV-2 genome sequencing by multiplex tiling PCR. PLoS ONE 15, (2020).

63. Nei, M. & Li, W.-H. Mathematical model for studying genetic variation in terms of restriction endonucleases (molecular evolution/mitochondrial DNA/nucleotide diversity). Genetics 76, 5269– 5273 (1979).

64. Been, F. et al. Population normalization with ammonium in wastewater-based epidemiology: Application to illicit drug monitoring. Environmental Science and Technology 48, 8162–8169 (2014).

65. Cori, A., Ferguson, N. M., Fraser, C. & Cauchemez, S. A new framework and software to estimate time-varying reproduction numbers during epidemics. American Journal of Epidemiology 178, 1505–1512 (2013).

66. Campbell, F. et al. Increased transmissibility and global spread of SARSCoV-2 variants of concern as at June 2021. Eurosurveillance 26, 1–6 (2021).

67. Davies, N. G. et al. Estimated transmissibility and impact of SARS-CoV-2 lineage B.1.1.7 in England. Science 372, (2021).

68. Washington, N. L. et al. Emergence and rapid transmission of SARS-CoV-2 B.1.1.7 in the United States. Cell 184, 2587-2594.e7 (2021).

69. Karthikeyan, S. et al. Rapid, Large-Scale Wastewater Surveillance and Automated Reporting System Enable Early Detection of Nearly 85% of COVID-19 Cases on a University Campus. 6, 793–814 (2021).

70. Calderón-Franco, D., Orschler, L., Lackner, S., Agrawal, S. & Weissbrodt, D. G. Monitoring SARS-CoV-2 in sewage: Toward sentinels with analytical accuracy. Science of the Total Environment 804, (2022).

71. Chan, M. C. W. et al. Seasonal influenza a virus in feces of hospitalized adults. Emerging Infectious Diseases 17, 2038–2042 (2011).

72. Pogka, V. et al. Laboratory surveillance of polio and other enteroviruses in high-risk populations and environmental samples. Applied and Environmental Microbiology 83, (2017).

73. Hong, P. Y. et al. Estimating the minimum number of SARS-CoV-2 infected cases needed to detect viral RNA in wastewater: To what extent of the outbreak can surveillance of wastewater tell us? Environmental Research 195, (2021).

74. Lynch, M., Bost, D., Wilson, S., Maruki, T. & Harrison, S. Population-genetic inference from pooled-sequencing data. Genome Biology and Evolution 6, 1210–1218 (2014).

75. Stern, A. et al. The unique evolutionary dynamics of the SARS-CoV-2 Delta variant-2 sequencing. doi:10.1101/2021.08.05.21261642.

76. Melnick, J. L. Poliomyekitis virus in urban sewage in epidemic and in nonepidemic times. American Journal of Epidemiology 45, 240–253 (1947).

77. Ye, Y., Ellenberg, R. M., Graham, K. E. & Wigginton, K. R. Survivability, Partitioning, and Recovery of Enveloped Viruses in Untreated Municipal Wastewater. Environmental Science and Technology 50, 5077–5085 (2016).

78. Wu, F. et al. SARS-CoV-2 Titers in Wastewater Are Higher than Expected from Clinically Confirmed Cases. mSystems 5, (2020).

79. Research Use Only 2019-Novel Coronavirus (2019-nCoV) Real-time RT-PCR Primers and Probes. CDC (2020).

80. Markt, R. et al. Detection and stability of SARS-CoV-2 fragments in wastewater: Impact of storage temperature. Pathogens 10, (2021).

81. Bushnell, B., Rood, J. & Singer, E. BBMerge – Accurate paired shotgun read merging via overlap. PLoS ONE 12, (2017).

82. Li, H. & Durbin, R. Fast and accurate short read alignment with Burrows-Wheeler transform. Bioinformatics 25, 1754–1760 (2009).

83. Grubaugh, N. D. et al. An amplicon-based sequencing framework for accurately measuring intrahost virus diversity using PrimalSeq and iVar. Genome Biology 20, (2019).

84. Wilm, A. et al. LoFreq: A sequence-quality aware, ultra-sensitive variant caller for uncovering cell-population heterogeneity from high-throughput sequencing datasets. Nucleic Acids Research 40, 11189–11201 (2012).

85. Li, H. A statistical framework for SNP calling, mutation discovery, association mapping and population genetical parameter estimation from sequencing data. Bioinformatics 27, 2987–2993 (2011).

86. Cingolani, P. et al. A program for annotating and predicting the effects of single nucleotide polymorphisms, SnpEff: SNPs in the genome of Drosophila melanogaster strain w1118; iso-2; iso-3. Fly 6, 80–92 (2012).

87. Cingolani, P. et al. Using Drosophila melanogaster as a model for genotoxic chemical mutational studies with a new program, SnpSift. Frontiers in Genetics 3, (2012).

88. Supplemental Material for A Better Lemon Squeezer? Maximum-Likelihood Regression With Beta-Distributed Dependent Variables. Psychological Methods (2006) doi:10.1037/1082-989x.11.1.54.supp.

89. Rigby, R. A. & Stasinopoulos, D. M. Generalized additive models for location, scale and shape. Appl. Statist vol. 54 (2005).

90. Barndorff-Nielsen, E. & Jorgensen, B. Some Parametric Models on the Simplex. Journal of multivariate analysis 39, 106–116 (1991).

91. Lee S, Wolberg G & Shin S.Y. Scattered data interpolation with multilevel B-splines. IEEE Transactions on Visualization and Computer Graphics 3, (1997).

92. Wastewater-based estimation of the effective reproductive number of SARS-CoV-2. doi:10.1101/2021.04.29.21255961.

93. Richter L, Trauner F, Schmid D & Stadlober E. Aktuelle Schätzung des seriellen Intervalles von COVID19, 2021, Österreich. https://www.ages.at/download/0/0/840fb2b85eb49b8f744d8d971614125ae197ab10/fileadmin/AGES2015/Wissen-Aktuell/COVID19/serial_interval_update2021_2021-06-14.pdf (2021).

94. Özkan, E. et al. High-throughput Mutational Surveillance of the SARS-CoV-2 Spike Gene. doi:10.1101/2021.07.22.21259587.

95. Nelson, C. W., Moncla, L. H. & Hughes, A. L. SNPGenie: Estimating evolutionary parameters to detect natural selection using pooled next-generation sequencing data. Bioinformatics 31, 3709– 3711 (2015).

96. Heiler, G. et al. Country-wide Mobility Changes Observed Using Mobile Phone Data during COVID-19 Pandemic. in Proceedings - 2020 IEEE International Conference on Big Data, Big Data 2020 3123–3132 (Institute of Electrical and Electronics Engineers Inc., 2020). doi:10.1109/BigData50022.2020.9378374.

